# A Working Model to Inform Risk-Based Back to Work Strategies

**DOI:** 10.1101/2020.12.18.20248512

**Authors:** Kristen Meier, Kirsten J. Curnow, Darcy Vavrek, John Moon, Kyle Farh, Martin Chian, Robert Ragusa, Eileen de Feo, Phillip G. Febbo

## Abstract

**Background:** The coronavirus disease 2019 (COVID-19) pandemic has forced many businesses to close or move to remote work to reduce the potential spread of disease. Employers desiring a return to onsite work want to understand their risk for having an infected employee on site and how best to mitigate this risk. Here, we modelled a range of key metrics to help inform return to work policies and procedures, including evaluating the benefit and optimal design of a SARS-CoV-2 employee screening program.

**Methods:** We modeled a range of input variables including prevalence of COVID-19, time infected, number of employees, test sensitivity and specificity, test turnaround time, number of times tested within the infectious period, and sample pooling. We modeled the impact of these input variables on several output variables: number of healthy employees; number of infected employees; number of test positive and test negative employees; number of true positive, false positive, true negative, and false negative employees; positive and negative predictive values; and time an infected, potentially contagious employee is on site.

**Results:** We show that an employee screening program can reduce the risk for onsite transmission across different prevalence values and group sizes. For example, at a pre-test asymptomatic community prevalence of 0.5% (5 in 1000) with an employee group size of 500, the risk for at least one infected employee on site is 91.8%, with 3 asymptomatic infected employees predicted within those 500 employees. Implementing a SARS-CoV-2 baseline screen with an 80% sensitivity and 99.5% specificity would reduce the risk of at least one infected employee on site to 39.4% and the predicted number of infected employees onsite (false negatives) to 1. Repetitive testing is required for ongoing vigilance of onsite employees. The expected number of days an infected employee is on site depends on test sensitivity, testing interval, and turnaround time. If the test interval is longer than the infectious period (∼14 days for COVID-19), testing will not detect the infected employee. Sample pooling reduces the number of tests performed, thereby reducing testing costs. However, the pooling methodology (eg, 1-stage vs 2-stage pooling, pool size) will impact the number of employees that screen positive, thereby affected the number of employees eligible to return to onsite work.

**Conclusions:** The modeling presented here can be used to help employers understand their risk for having an infected employee on site. Further, it details how an employee screening program can reduce this risk and shows how screening performance and frequency impact the effectiveness of a screening program. The primary factors determining the effectiveness of a screening program are test sensitivity and frequency of testing.

**Disclaimer:** This publication is offered to businesses/employers as a model of potential risk arising from COVID19 in the workplace. While believed to be based on reliable data, the model described herein has not been prospectively validated and should not be relied upon for any purpose other than as an aid to understand the potential impacts of a number of variables on the risk of having COVID19 positive employees on a worksite. Decisions related to workplace safety; COVID19 related workplace testing; programs and procedures should be based upon your actual data and applicable laws and public health orders.

## INTRODUCTION

The coronavirus disease 2019 (COVID-19) pandemic has had a dramatic impact on our global health and economy. As of Dec 4, 2020, there have been over 64 million confirmed cases and 1.5 million deaths globally and the numbers continue to climb.^1^ In order to reduce the toll on human health, countries and municipalities have embraced social distancing measures to reduce community-based transmission. These measures forced some non-essential businesses to close and others to dramatically change their operations. While there is growing evidence that these measures have reduced the reproductive factor (the “R_0_”) of SARS-CoV-2, the virus that causes COVID-19, they have also had a dramatic impact on our global, local, and family economies.

As transmission was reduced and controlled in some communities, there was increasing focus on a safe return to work to re-establish economies. However, COVID-19 remains in most communities, with data from Hong Kong^2^, China^3^, and Singapore^4^ showing continued vigilance is required to avoid large clusters from arising and second waves. There are a growing number of voices suggesting that broad-based testing is required to keep numbers under control. Broad-based testing may be of value to employers where a single infected employee could result in a site shut down.

Employers are now seeking to understand their optimal return to work strategy, such as leveraging SARS-CoV-2 testing and evaluating employees for signs and symptoms of COVID-19, in order to reduce the risk of an employee introducing COVID-19 to the workplace. The optimal strategy will be dependent on multiple characteristics of the virus, community, and the testing approach, in addition to the size of the organization and ability to put in place other measures such as social distancing. In an ideal world, everyone could be screened any time they came on site. The results would be fast and 100% certain. In practice, SARS-CoV-2 testing takes time, costs money, and is less than 100% accurate (sensitive and specific). Further, there are practical limits to how often testing can be performed and how fast results can be obtained, and there are many unknowns.

With the broad availability of diagnostic testing, we are getting a better understanding of community prevalence of SARS-CoV-2, which is critical to any individual’s risk of exposure, and some of the basic viral characteristics are becoming better known. The clinical course of viral infection exhibits broad ranges of severity and a significant proportion (∼30%) of confirmed patients remain asymptomatic.^5, 6^ It is estimated that the average incubation period between exposure and onset of symptoms is 5 days, with 98% of those that will eventually exhibit symptoms doing so within 12 days of exposure.^7^ Current reporting of the onset and duration of the infectious period suggests it may be quite variable between patients^8-10^. Further, as most available data on the infectious period is based on symptomatic patients, the transmissibility and infectious period in asymptomatic patients is even less certain^11^. Finally, with the clarity of the clinical course and reference method testing modality, the analytic and clinical performance of testing (e.g., sensitivity, specificity) is better defined.

There are some general concepts that help in understanding the benefits and limitations of screening. Screening is effective and useful when the disease prevalence is not too low (if no one in the community has the disease, screening will have no impact) or too high (wide community spread, so screening has limited benefit). The goal of screening is to keep the number of accepted individuals under the maximum number considered acceptable and/or achievable. For example, an intuitively appealing goal would be to keep the max number of infected individuals in a group below 1, with the number of infected employees calculated as the Number of Employees x Employee prevalence. Employee prevalence is basically community prevalence unless some form of frequent screening is performed to isolate infected employees. Another important concept is that to reduce the prevalence in the employee group by screening, tests must be performed more frequently than the course of the disease. Thus, the interval between employee screens must be substantially shorter than the 14 to 21 day disease progression time^7, 8, 10-12^ to be effective; sampling at a time interval longer than once a week won’t be expected to have a substantial impact. For example, universities in the US that have implemented testing are sampling twice a week.^13, 14^ Other studies have shown that with measures that reduce the disease R0 to ∼1.5, testing once per week may be sufficient.^15^ However, as sampling is only ever going to be every day at best (and probably every other day), the most we can expect to be able to reduce employee prevalence using screening is by around 10 fold. That sets the upper bound of prevalence where no amount of screening will be effective at keeping Number of Employees x Employee prevalence below 1.

In this paper, we modelled a range of key metrics to enable development of a return to work policy for our employees. We evaluated factors such as prevalence, group size, screening performance, screening frequency, and sample pooling on the risk of an infected person coming on site (and therefore potentially transmitting the virus to other employees or group members). Using this information, we can estimate, for a given moment in time, the number of employees that would screen negative and could return to work and those that would screen positive and therefore not return to work (Figure 1). We leverage available literature – both peer-reviewed and pre-print articles – to provide ranges for each variable. In addition, we are currently developing an online version of a tool where a user can provide the number of employees, the pre-test prevalence of SARS-CoV-2, test performance (sensitivity and specificity) and their threshold of risk in order to explore different testing schedules for a safe return to work. The goal of the modeling presented here is to provide information that can be used to answer questions and inform decisions around employee screening programs including testing frequency, testing schemes, individual versus sample pooling, and test performance.

**Figure 1.**
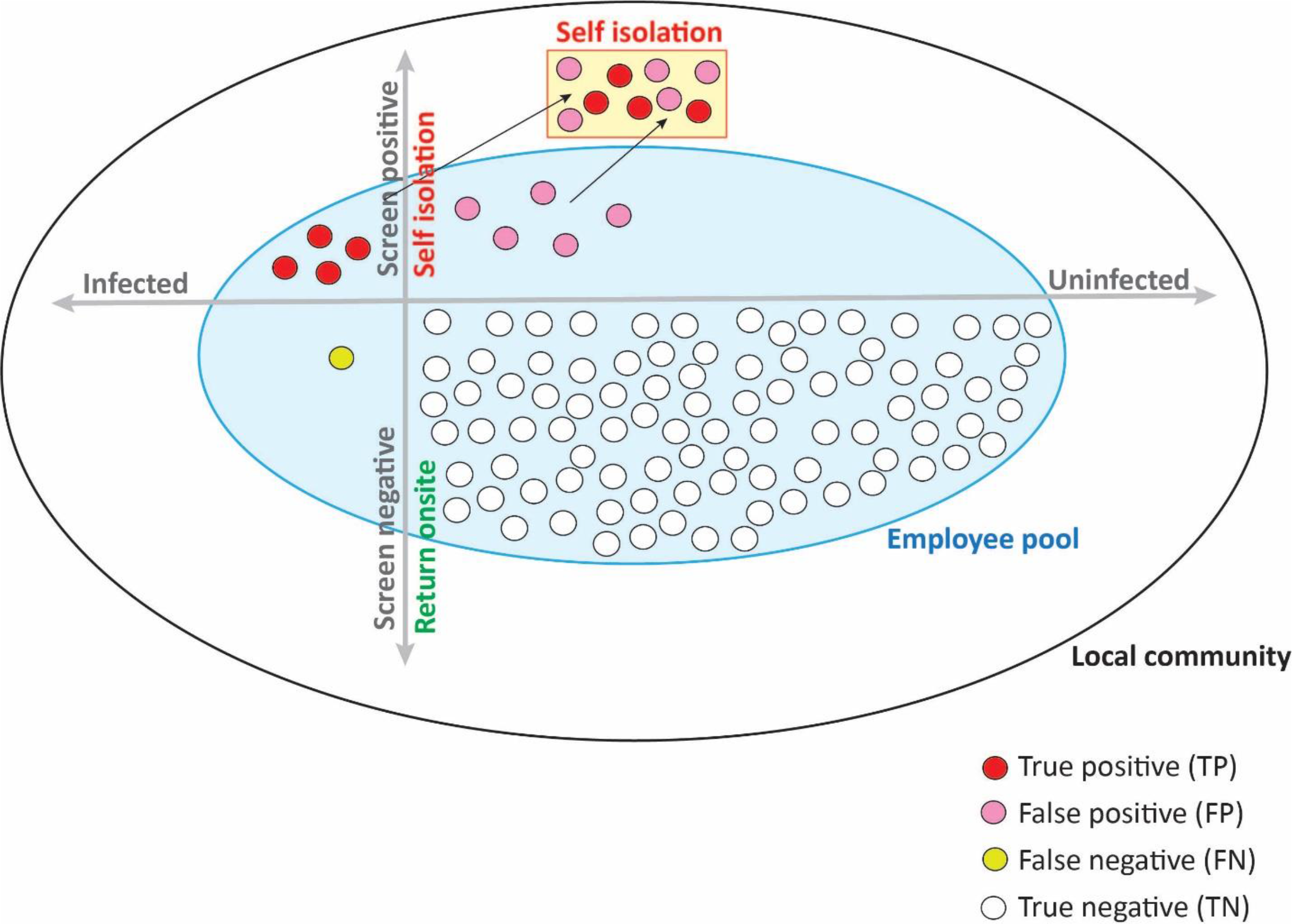
Return to work screening model. SARS-CoV-2 screening is performed for asymptomatic employees. Employees that screen negative are permitted to return to onsite work. Employees that screen positive go into self - isolation.

## METHODS

Models are provided for metrics that can enable decision makers to understand the implications of different screening options (different test performance, individual vs pooled samples, and testing frequency) under different scenarios (prevalence and employee group size). These models will help a user understand the following: 1, how many employees would screen positive versus negative and the likelihood of them being infected versus uninfected (positive predictive value, negative predictive value); 2, impact of test performance on the risk or probability of an infected employee arriving at work on site; 3, influence of group size on the risk of having any (at least one) infected person(s) on site; 4, implications of screening interval (eg, daily vs weekly screening) on the length of time an infected person is on site; and 5, how local prevalence rates can impact the decision on whether to screen or not. Screening approaches explored include symptom screening alone (no SARS-CoV-2 testing) and screening asymptomatic employees using SARS-CoV-2 testing, including testing before returning on site (baseline testing) and frequent onsite testing. All formulas used as part of the models are detailed in Supplementary Table 1.

**Table 1.** Definitions and ranges of model input variables

| Variable | Units | Definition | Range of Values | References |
| --- | --- | --- | --- | --- |
| P | percentage | Prevalence of COVID-19 in population of interest | 0.001%, 0.01%, 0.1%, 0.5%, 1.0%, 2.0% | 16, 50 |
| TINF | time | time infected, asymptomatic but contagious | 14 days, 21 days | 7, 8, 10-12 |
| NE | number | number of employees in a group that is screened for returning to work on site | 20, 50, 100, 250, 500, 1000 | N/A |
| se | percentage | sensitivity of test; probability that infected employee will have positive test result | 70%, 80%, 90% | 29 |
| sp | percentage | specificity of test; probability that uninfected employee will have negative test result | 98%, 99%, 99.5%, 99.9% | 27, 28 |
| TA | time | Turn-around time from sample collection to result | 1 or 2 days | N/A |
| TI | time | Testing interval (time between tests); $TI \geq TA$ | 1-14 days; $TI \geq TA$ | N/A |
| m | number | (Implicit) Number of times individual tested per time infected; $m \leq TINF/TI$ | 1-21 | N/A |
| k | number | specimen pool size | 5, 10 | N/A |
| se.k | percentage | sensitivity of test when using pooled specimens | se-1.5 for k=5<br>se-2.5 for k=10 | N/A |
| sp.k | percentage | specificity of test when using pooled specimens | sp for k=5 or 10 | N/A |

N/A, not applicable indicates no appropriate references were identified or not required for that variable.

### Model Inputs – Variable Definitions and Ranges

Model inputs may be uncontrolled or controlled to some degree by the employer. Some inputs such as community prevalence will differ across communities and change over time during the pandemic. Other inputs such as test performance (sensitivity and specificity) and turnaround time for a test result may be somewhat controlled in test development or by the choice of technology. The range of inputs considered in the model are described in Table 1 below. Where data were available to guide the ranges, references are indicated.

Prevalence will vary depending on the surrounding work community and will change over time. Current prevalence estimates may be difficult to obtain. Prevalence estimates can be made from the percentage of the population undergoing testing in a given (recent) period of time and the percentage of those that are positive for COVID-19. Nationwide in the US, the CDC reported 1.8M specimens tested with a positive rate of 8.6% for the week ending July 18^16^; as a proportion of the total US population (∼330M^17^), this is a positive rate of around 0.5%. Importantly, the accuracy of prevalence estimates will be impacted by the proportion of the population undergoing testing, and to what degree testing is targeted/restricted to symptomatic people. In the US during the summer of 2020, testing was largely limited to symptomatic individuals, and with around 30% of infected individuals are asymptomatic^5, 6^, the 8.6% positive rate above was unlikely to have captured all infected cases. Testing a larger overall proportion of the population, and including asymptomatic individuals, will provide more accurate values. Further, there is large regional variation in COVID-19 prevalence, so national numbers may not be the best indicator of local prevalence. As such, identifying a local source of information for testing numbers and positivity rates, such as state public health department websites^18, 19^ and county websites,^20, 21^ will help to more accurately predict prevalence in the local community encompassing the employee base. In addition, there are a number of online tools available to help guide estimation of local prevalence^22-25^. The infectious period is estimated to be around 14 to 21 days (maybe longer in some cases), with an onset at around a couple days after exposure^11, 26^. As such, we modelled time infected (TINF) values of 14 and 21 days.

The sensitivity and specificity of the test will depend on the SARS-CoV-2 screen technology, sample type, whether testing individual or pooled samples, and test development optimization (eg, threshold for positivity, sequencing depth)^27-31^. A pooled sample is one where individual samples are combined and the pool is tested rather than the individual samples. Test sensitivity for pooled samples may be reduced compared with individual samples since the viral RNA is diluted. It is assumed that the specificity would not change with test pooling. However, if the viral load in most samples is still above the test threshold, the sensitivity may not be greatly reduced by pooling. Empirical studies of pooling for SARS-CoV-2 have indicated that Ct (PCR cycle time) increases by approximately log2(pool size) as expected^32^; as such pooled testing of 5 samples would increase Ct by an expected value of 2.3. Based on empirical distributions of Ct values, we estimate a decrease in sensitivity of 1.5% if the Ct value of all individuals is shifted by 2.3 (pooling n=5) and assuming a threshold of detection of Ct=40, and 2.5% if the Ct was shifted by 3.3 (pooling n=10).^33^

Some tests allow for an indeterminate or equivocal result that is valid (passes quality control metrics) but is not declared positive or negative. If indeterminate results are excluded from reported test performance (sensitivity and specificity) estimates, the in-use test sensitivity and specificity may be lower than what is reported, so lower values should be considered in the model.

### Models and Model Outputs

The testing schemes modelled included: Symptom screening alone (no SARS-CoV-2 testing); baseline SARS-CoV-2 screening on asymptomatic employees; and repeated SARS-CoV-2 testing. Models for testing using an imperfect test (sensitivity and specificity < 100%) are described for individual samples and pooled samples. Different approaches can be taken to manage test results. Here, employee management was as follows: Test negative, OK to return onsite work; test positive, self-isolation with or without follow-up testing. Model outputs calculated are described in Table 2. All formulas used in this modelling are detailed in Supplementary Table 1.

**Table 2.** Definition of model output variables and typical values

| **Variable** | **Units** | **Definition** |
| --- | --- | --- |
| NI | number | number of infected employees out of NE screened at point in time |
| NH | number | number of “healthy” employees, where healthy means uninfected with respect to SARS-CoV-2 |
| Npos | number | number of test positive employees |
| Nneg | number | number of test negative employees |
| NQ | number | number of employees quarantined |
| TP | number | Number of employees with true positive test result; employee is infected and has positive test result |
| FP | number | Number of employees with false positive test result; employee is uninfected and has positive test result |
| FN | number | Number of employees with false negative test result; employee is infected and has negative test result |
| TN | number | Number of employees with true negative test result; employee is uninfected and has negative test result |
| PPV | Percent | positive predictive value; posttest probability that a person who is screen positive is infected |
| 1–PPV | Percent | posttest probability that a person who is screen positive is uninfected/healthy |
| NPV | Percent | negative predictive value; posttest probability that a person who is screen negative is uninfected/healthy |
| 1–NPV | Percent | posttest probability that a person who is screen negative is infected |
| TC | Time<br>(day) | Time an infected employee is contagious, on site |

### No SARS-CoV-2 testing - individual samples

When no testing is performed prior to an employee returning to work, the expected number of infected employees out of the number of employees (NE) returning on site is calculated as the number of employees times prevalence. If employees are prescreened for signs and symptoms of infection (eg, thermal scanning), then it is the prevalence among asymptomatic individuals prescreened as negative. The risk of having any infected employee on site is the probability that at least 1 employee out of NE employees on site is infected (Supplementary Table 1, Formula 1).

### Baseline SARS-CoV-2 testing - individual samples

An employer may consider using SARS-CoV-2 testing as a baseline screen for a group of NE asymptomatic employees for return to work on site. Some of these employees may be infected (with probability equal to prevalence), and others will be uninfected or “healthy” with respect to SARS-CoV-2. Those who test negative may return to work on site while those who test positive are required to isolate at home. Most of the healthy employees will test negative or have a true negative [TN] result with probability equal to specificity; most of the infected employees will test positive or have a true positive [TP] result with probability equal to sensitivity. A small percentage of the infected employees will test negative or have a false negative [FN] result; and, a small percentage of the healthy employees test positive or have a false positive [FP] result. The possible outcomes are depicted in Figure 1.

The expected number of employees with a given result will depend on the number of employees, prevalence, and the test sensitivity and specificity. The cross tabulation of expected test results versus true status can be displayed as shown in Supplementary Table 2 and are calculated using the set of formulas in No. 2, Supplementary Table 1.

The post-test likelihood of a screen negative employee being uninfected (negative predictive value of the screen) and the post-test likelihood of a screen positive employee being infected (positive predictive value of the screen) are calculated using Formulas 3 and 4, respectively, in Supplementary Table 1. The post-test risk of having any infected employee on site is the probability that at least 1 employee out of test negative employees on site is infected, that is, has a false negative result. This risk was calculated using Formula 5 in Supplementary Table 1.

### Baseline SARS-CoV-2 testing - pooled samples

Pooled testing, also known as group testing^34^, is a procedure that that can be used when prevalence is small in order to reduce the number of tests performed^35, 36^. A simple version of this procedure was proposed by Dorfman^37^ to screen US soldiers for syphilis during World War II. The basic idea of this 2-stage approach is to combine individual samples into pools of size k (eg, 5 or 10) and test each pool. For a pooled sample, the assumed impact on test sensitivity is summarized in Supplementary Table 1 – Formula 6. If the pool is test negative, then everyone in the pool is declared test negative. If the pool is test positive, then leftover sample from each of the k individuals in the pool are tested separately and the declared test result is based on the individual test. A simplified (1-stage) version of this approach is to use the pooled result as the declared result for each individual. The impact of the Dorfman (2-stage) and simplified (1-stage) testing approaches on the probability of an infected employee on site as well as the expected number of positive results was evaluated. The risk of having any infected employee on site using the 1-stage and 2-stage pooled sample approach is calculated by simulation assuming individual outcomes are independent Bernoulli trials.

Calculation of the risk of an infected employee on site based on a pooled sample approach utilizes the prevalence of infected pools or pre-test probability that a pool is infected rather than the pre-test probability that an individual is infected. A pool is infected if at least 1 sample in the pool is from an infected employee (Supplementary Table 1 – Formula 7).

### Repeated testing - individual samples

The ability of testing to detect an infected employee arriving at work depends on test sensitivity and number of tests performed. The time between tests (testing interval) and turnaround time from sample collection to result will determine the expected number of days an infected employee will be on site before being detected.

The probability that an infected employee will be detected by each test cycle is calculated from test sensitivity as described in Supplementary Table 3. The expected number of days an infected employee will be on site before being detected additionally depends on the test interval and turnaround time (Supplementary Table 1 – Formula 8, and Supplementary Figure 1). The intuition for this calculation is depicted in Supplementary Figure 2. These formulas assume that each testing event is independent, and the employees test result (positive or not) follows a Bernoulli distribution with probability sensitivity.

**Table 3.** Impact of the prevalence of infected individuals on prevalence of infected pools*

| Prevalence of infected individual | Prevalence across pools by number of samples (k) in pool |  |  |  |  |
| --- | --- | --- | --- | --- | --- |
|  | k=2 | k=5 | k=10 | k=15 | k=20 |
| **0.001%** | 0.002 | 0.005 | 0.010 | 0.015 | 0.020 |
| **0.01%** | 0.02 | 0.05 | 0.10 | 0.15 | 0.20 |
| **0.1 %** | 0.2 | 0.5 | 1.0 | 1.5 | 2.0 |
| **1 %** | 2 | 5 | 10 | 14 | 18 |
| **5 %** | 10 | 23 | 40 | 54 | 64 |
| **10%** | 19 | 41 | 65 | 79 | 88 |

\* Prevalence of infected pools is the probability that a pool is infected. A pool is infected if at least one sample in the pool is from an infected individual.

**Figure 2.**
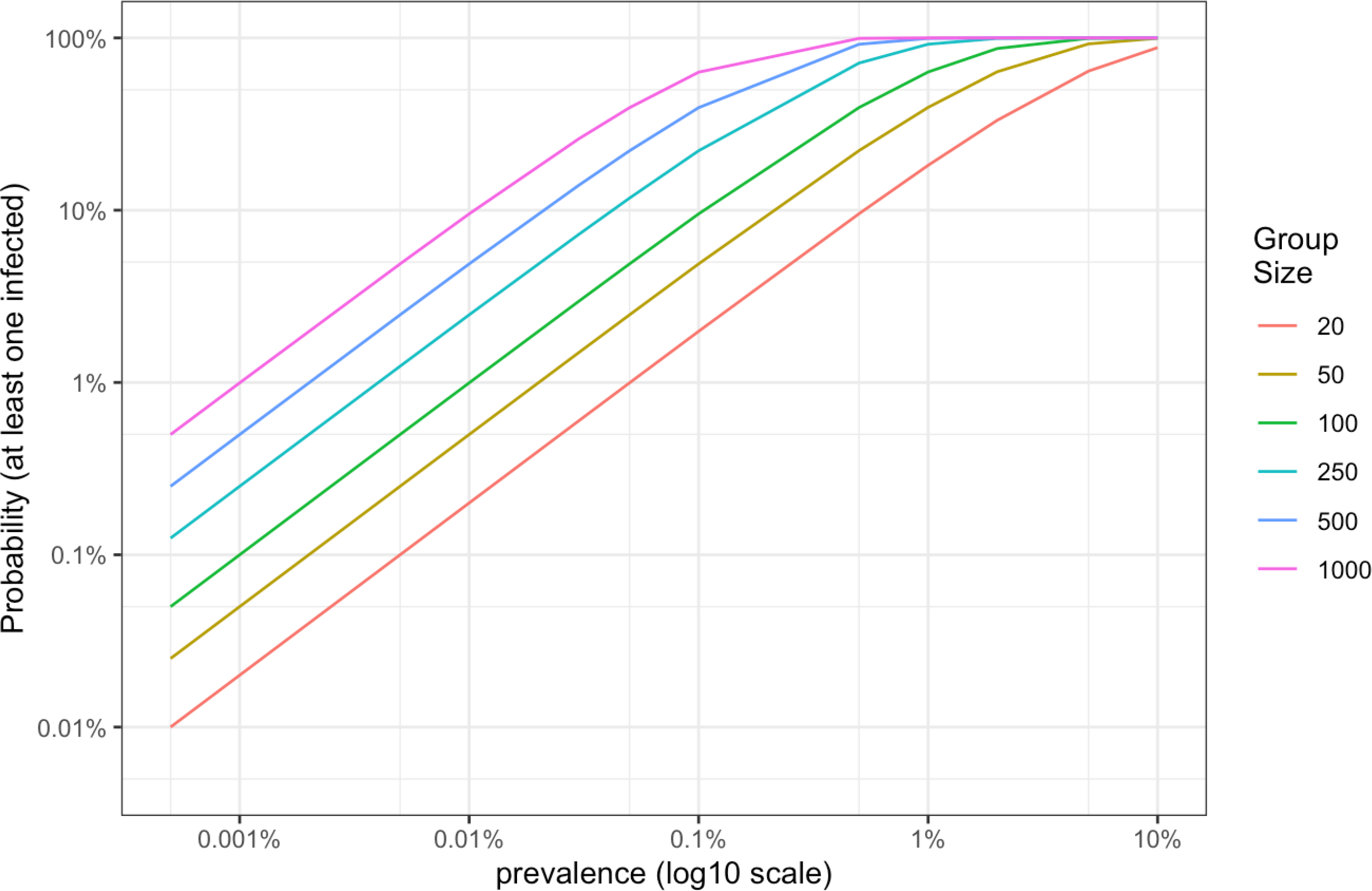
Probability of having at least one infected employee on site for different employee group sizes and at different prevalences in the absence of an employee screening program.

## RESULTS

### No screening modelling

Without any screening, the chance that an infected employee arrives at work on site is equal to the community prevalence of COVID-19. If employees are screened for temperature or signs and symptoms, then the COVID-19 prevalence in a group of employees may be reduced. The implementation of social distancing and use of personal protective equipment (PPE) such as masks can reduce the chance of transmission should an affected employee return on site. We modelled the probability of having at least one infected employee on site in the absence of a broad screening program at different prevalences and for different employee group sizes. Figure 2 shows that as prevalence and group size increase, there is a higher probability of having at least one infected employee on site. At a prevalence of 0.1% (or 1 in 1000) and 20 employees, there is a 2% probability of having at least one infected employee on site in the absence of an employee screening program. However, if the prevalence in asymptomatic people is 0.5% and 1000 employees come on site, the probability or risk of having at least one infected employee on site is 99.3%. In this second example, the expected or average number of infected employees on site is 5. These 5 asymptotic employees would remain on site until signs and symptom appear or the infection resolves, potentially transmitting the virus to other employees on site. Without testing or symptom screening, the only way an employer can reduce the chance of an infected employee returning on site is to limit the group size of employees returning on site.

### Baseline Employee Screening to Reduce Risk before Returning to Work

One way to reduce the risk of workplace transmission is to screen employees using a SARS-CoV-2 test before returning to work. This would reduce the workplace prevalence when employees first return on site. Figure 3 shows the post-test workplace prevalence as a function of (pre-test) prevalence and test sensitivity, including a dashed reference at no screening for easy comparison. At a prevalence of 0.1%, when test sensitivity is 80% (olive line), the post-test prevalence is reduced to 0.02%, a 5-fold reduction in prevalence (vertical distance between the pink dashed line and the olive line). If test sensitivity is higher at 90% (green line) the reduction is greater; the post-test prevalence is reduced to 0.01%, a 10-fold reduction in prevalence. Expected post-test prevalences depicted in Figure 3 are provided in Supplementary Tables 4-7 for sensitivities 70%, 80%, 90%, and 95%, respectively, under “prevalence in screen negatives (1− NPV).

**Table 4.** Probability an infected employee will be detected by each test cycle

| Test Cycle* | 70% Sens | 80% Sens | 90% Sens | 95% Sens |
| --- | --- | --- | --- | --- |
| 1 | 70.0% | 80.0% | 90.0% | 95.0% |
| 2 | 91.0% | 96.0% | 99.0% | 99.8% |
| 3 | 97.3% | 99.2% | 99.9% | 100.0% |
| 4 | 99.2% | 99.8% | 100.0% | 100.0% |
| 5 | 99.8% | 100.0% | 100.0% | 100.0% |

\*Test cycles may consist of any combination of pre-onsite and once onsite testing; Sens, sensitivity.

**Figure 3.**
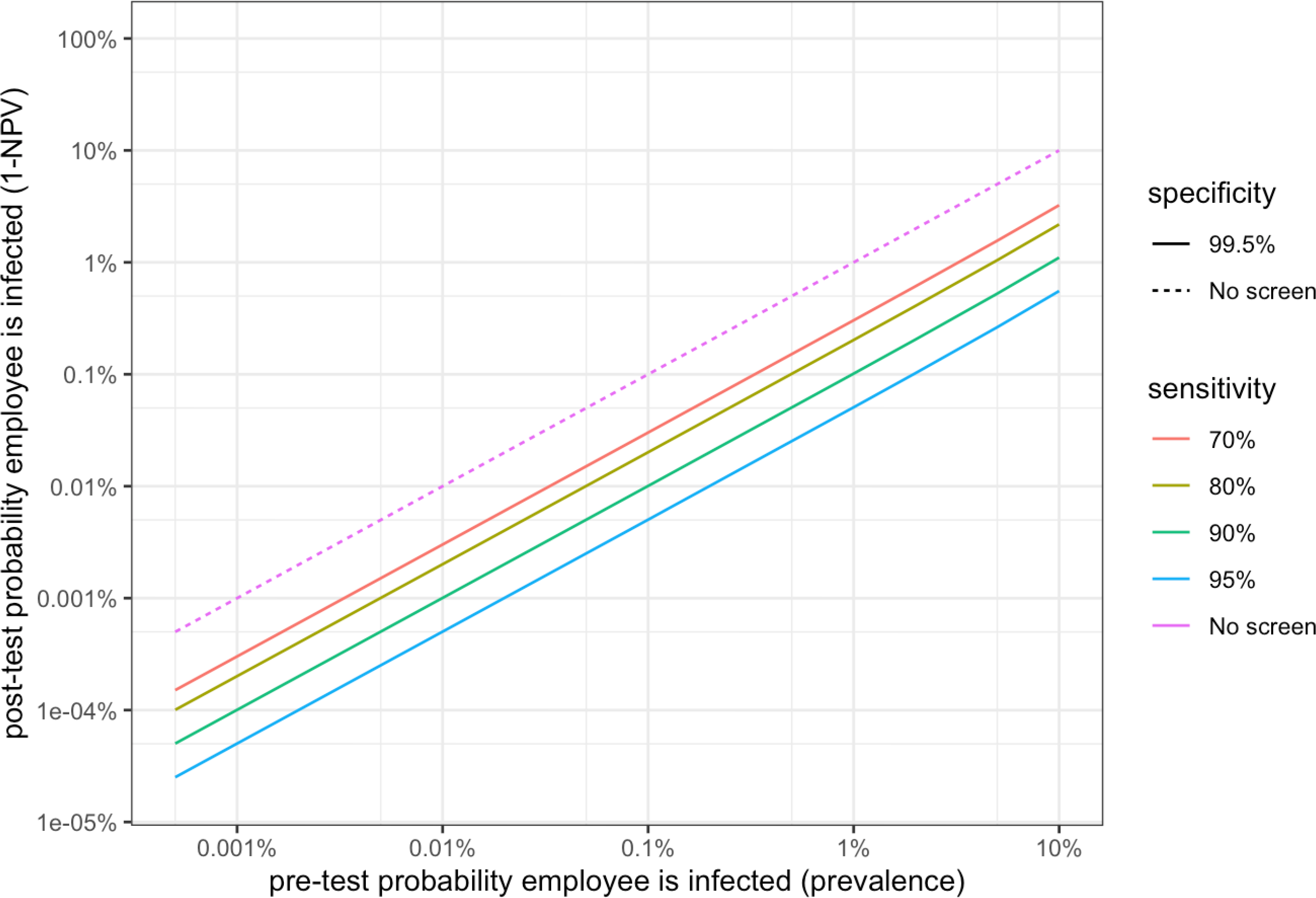
Probability that a single screen negative employee is infected with COVID-19 [false negative result] versus log10(prevalence) for a range of sensitivity values at set specificity of 99.5%.

To understand the impact of the workplace prevalence reduction, we next modelled the risk of at least one infected employee (a false negative) on site after implementation of a baseline employee screening program. We used fixed test performance values of 80% sensitivity and 99.5% specificity. The expected number of screen negatives and screen positives using this screening example is shown in Supplementary Table 5. In this model, screen positive employees would stay home while screen negative employees would return to onsite work. Figure 4 shows the probability of having at least one infected employee on site with the introduction of a baseline employee screening across three prevalences and a range of group sizes. With this example baseline test, the introduction of an employee screening program would reduce the probability on having at least one infected employee on site across group sizes and prevalences (Figure 4, Figure 5).

**Table 5.** Cross tabulation of expected SARS-CoV-2 Test Result versus the true status in 1000 employees tested based on 0.5% (5:1000) prevalence, 80% test sensitivity, and 99.5% test specificity

|  |  | True Status |  | Total |
| --- | --- | --- | --- | --- |
|  |  | Infected | Uninfected<br>"Healthy" |  |
| SARS-CoV-2<br>Test Result | Test Positive | 4 | 5 | 9 |
|  | Test Negative | 1 | 990 | 991 |
| Total |  | 5 | 995 | 1000 |

**Figure 4.**
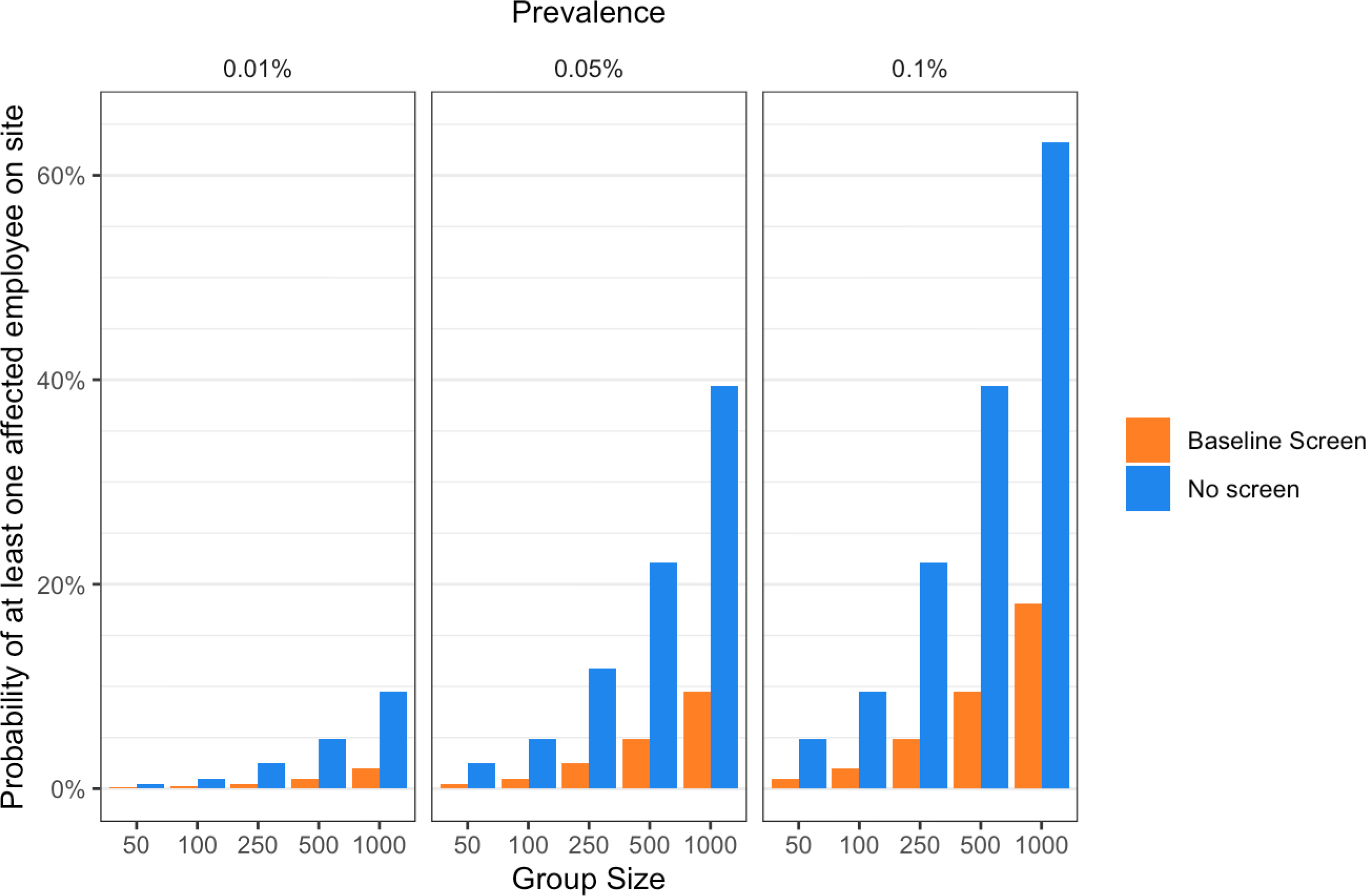
Impact of baseline screening on probability of an infected employee being on site.

**Figure 5.**
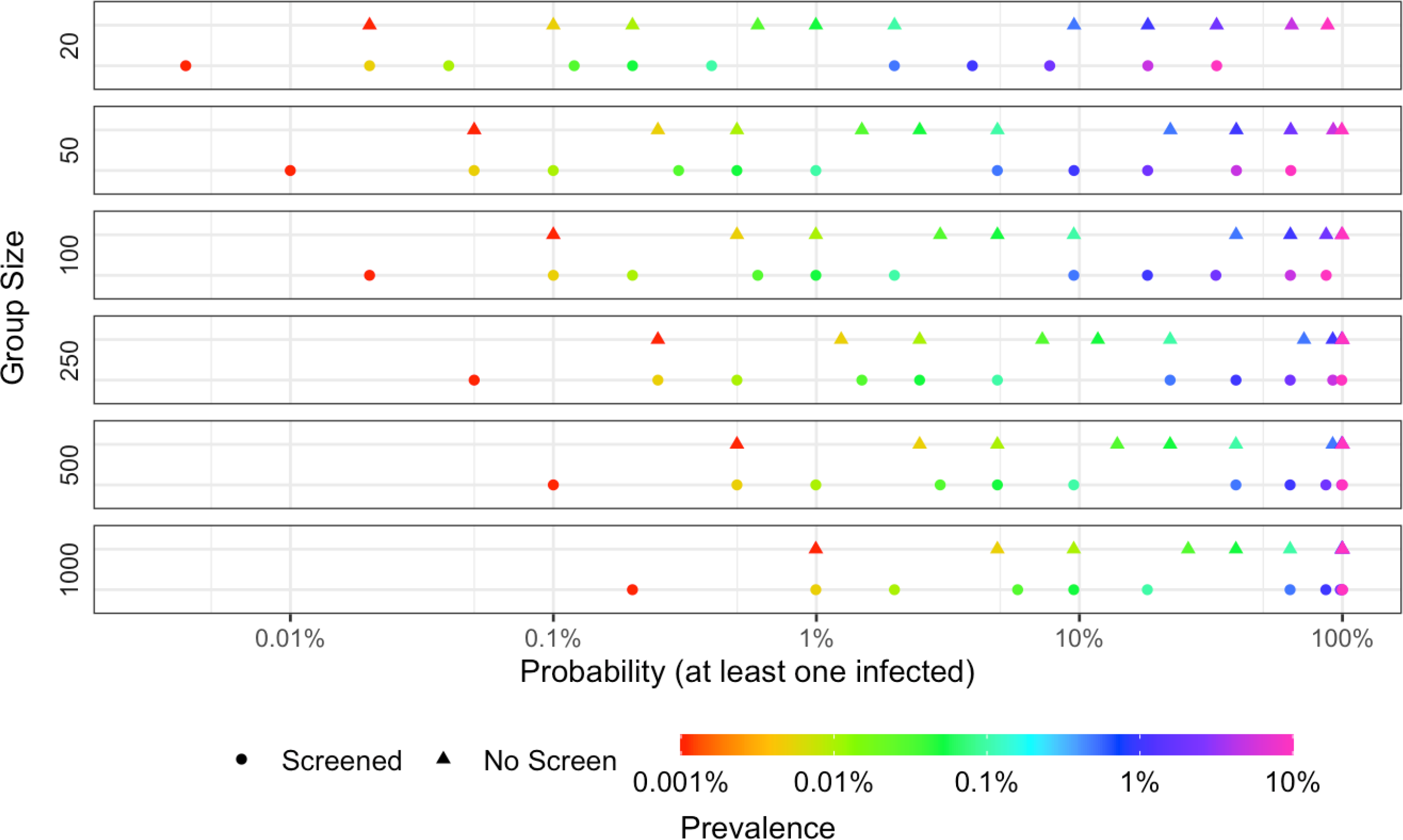
Probability of at least one infected screen negative employee based on a single screen of 80% sensitivity and 99.5% specificity.

For example, at a pre-test community prevalence of 0.01% (or 1 in 10,000), the baseline screening has reduced the prevalence in the workplace population that would return to work (screen negative) to 0.002%, an absolute difference of 0.008% (Supplementary Table 5). That is, for an employee that is screen negative, there is a 0.002% chance they are infected (a false negative). When applied to an employer testing 1000 employees for return to work (0.01% prevalence; 80% sensitivity, 99.5% specificity), there would be around 995 test negative employees that would return to work. There is a 2.0% chance that at least one of the test negative employees is infected (false negative) and the expected or average number of infected employees on site is <1 employee. The chance of an infected employee returning to work has been reduced from 9.52% (no testing) to 2.0% with a baseline screen, an absolute difference of 7.52% (=9.52-2.0) or fold difference of 4.8 (9.52/2.0). However, if an employer’s risk threshold for having at least one infected employee on site is 1%, then an employee group size of 1000 would not be acceptable (2.0% risk), whereas a group of 250 employees for return to work (0.5% risk) may be acceptable.

### Impact of Screening Test Performance

Next, we modeled the impact of test performance on a baseline employee screening program. For the narrow range of specificities considered, specificity has virtually no impact on the probability of a screen negative employee truly being infected (Figure 6); however, specificity will impact the number of false positives, and therefore the number of employees that would be asked to stay home unnecessarily. Subsequent results will consider a single specificity of 99.5%. In contrast, test sensitivity has a marked impact on the post-test probability a single screen negative employee is infected (false negative). A more sensitive test will also reduce the risk of having an infected employee return on site. With an employer’s risk tolerance for having at least one infected employee returning to work on the y-axis, and if prevalence is <0.01% then an employee group size of 20 employees on site may be acceptable even when test sensitivity is as low as 70%. If the group size is 500, even a test with 95% sensitivity would not meet the employer’s needs. Values for a range of prevalence values and group sizes are provided for 70% (Supplementary Table 4), 80% (Supplementary Table 5), 90% (Supplementary Table 6), and 95% (Supplementary Table 7) sensitivity. A 10% increase in sensitivity from 80% (Supplementary Table 5) to 90% (Supplementary Table 6) results in a halving of the post-test prevalence in screen negative employees.

**Figure 6.**
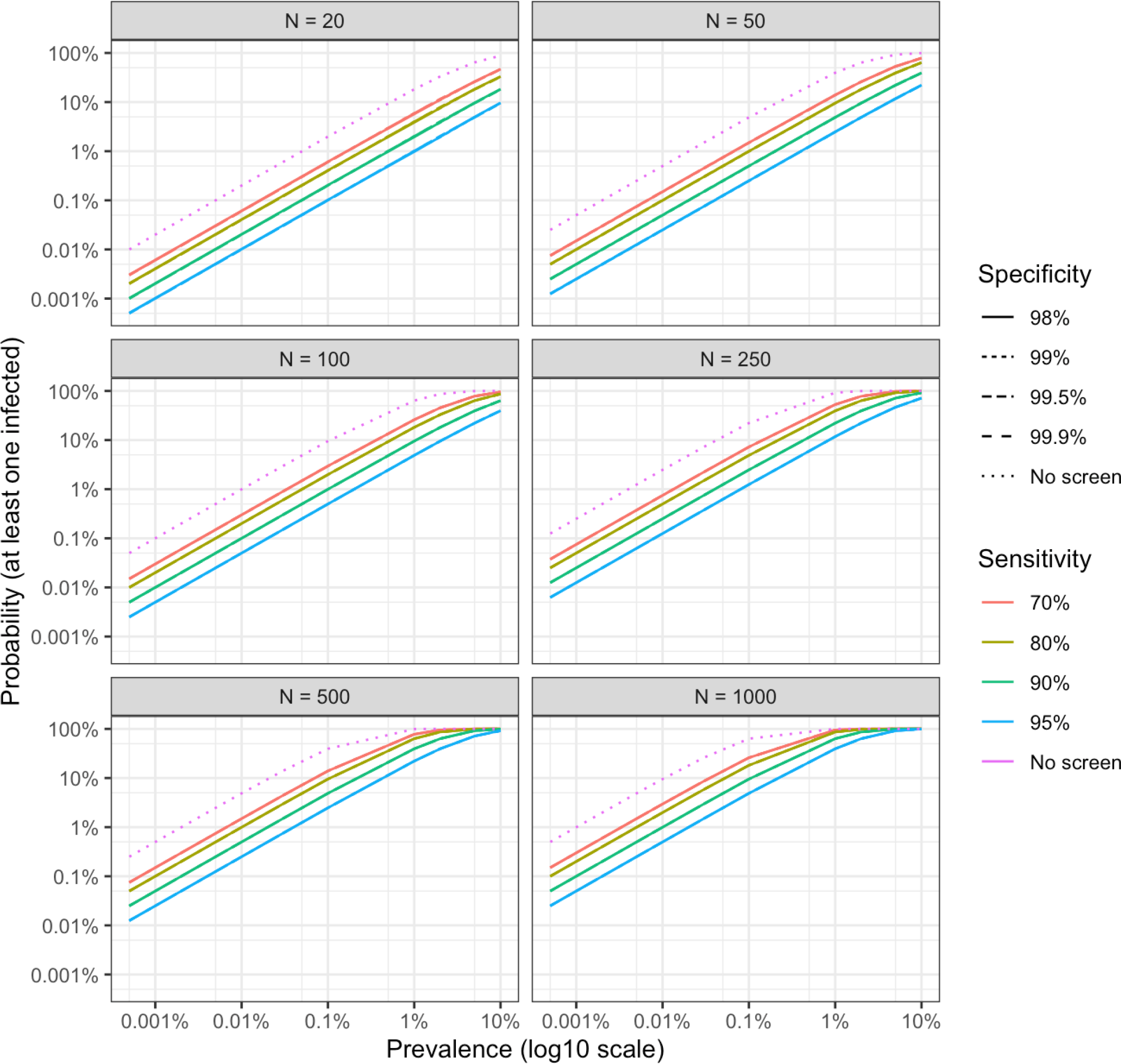
Probability that at least one screen negative employee returning on site is infected with COVID-19 [false negative result] versus log10(prevalence) for a range of sensitivity and specificity values. Specificity lines are overlapping for each given sensitivity and thus not visible.

### Impact of Sample Pooling on Screening Performance

The expected number of infected employees on site and the probability of an infected employee on site were estimated for both the 2-stage Dorfman pooled testing approach and a 1-stage pooled approach (pooled result as the declared result for each individual) for baseline testing. The expected prevalence of infected pools is greater than the expected prevalence of infected individuals roughly by a factor of pool size as shown in Table 3. Pooled testing reduces the number of tests performed, thereby reducing testing costs. However, it is important to also consider the impact on the number of employees that screen positive and the probability of an infected employee on site. The chance of an infected employee on site under baseline 1-stage testing (Supplementary Table 8) is less than that under 2-stage testing (Supplementary Table 9), but the number of false positives is higher. The 1-stage pooled approach will have a significant impact on the number of employees that screen positive (increasing by a factor of the pool size), and as such, will reduce the number of employees that return to work. Thus, the cost savings of pooling should be balanced against the impact on productivity in terms of the number of employees able to return on site.

### Impact of Repetitive Testing and Testing Interval (time an infected employee remains on site)

While baseline testing can reduce the initial risk on an infected individual returning to work, until the community prevalence drops substantially or a vaccine is available, there remains a significant risk for employees to become infected through community transmission. As such, repetitive testing is required for ongoing vigilance of onsite employees. The ability of testing to detect an infected employee arriving at work depends on test sensitivity and number of tests performed. The time between tests (testing interval) and turnaround time to a result will determine the expected number of days an infected employee will be on site before being detected (Figure 7). Table 4 shows how test sensitivity impacts the chance of detecting an infected employee as a function of number of tests performed. If the infected contagious employee on site is tested twice during the infectious period using a test with 80% sensitivity, there is an 80% chance that the employee will be test positive by Test 1 and a 96.0% chance they will test positive by Test 2. If the employee had been tested once prior to coming on site, the chance they would arrive on site and be detected by the second test on site (3rd test cycle) is 99.2%.

**Figure 7.**
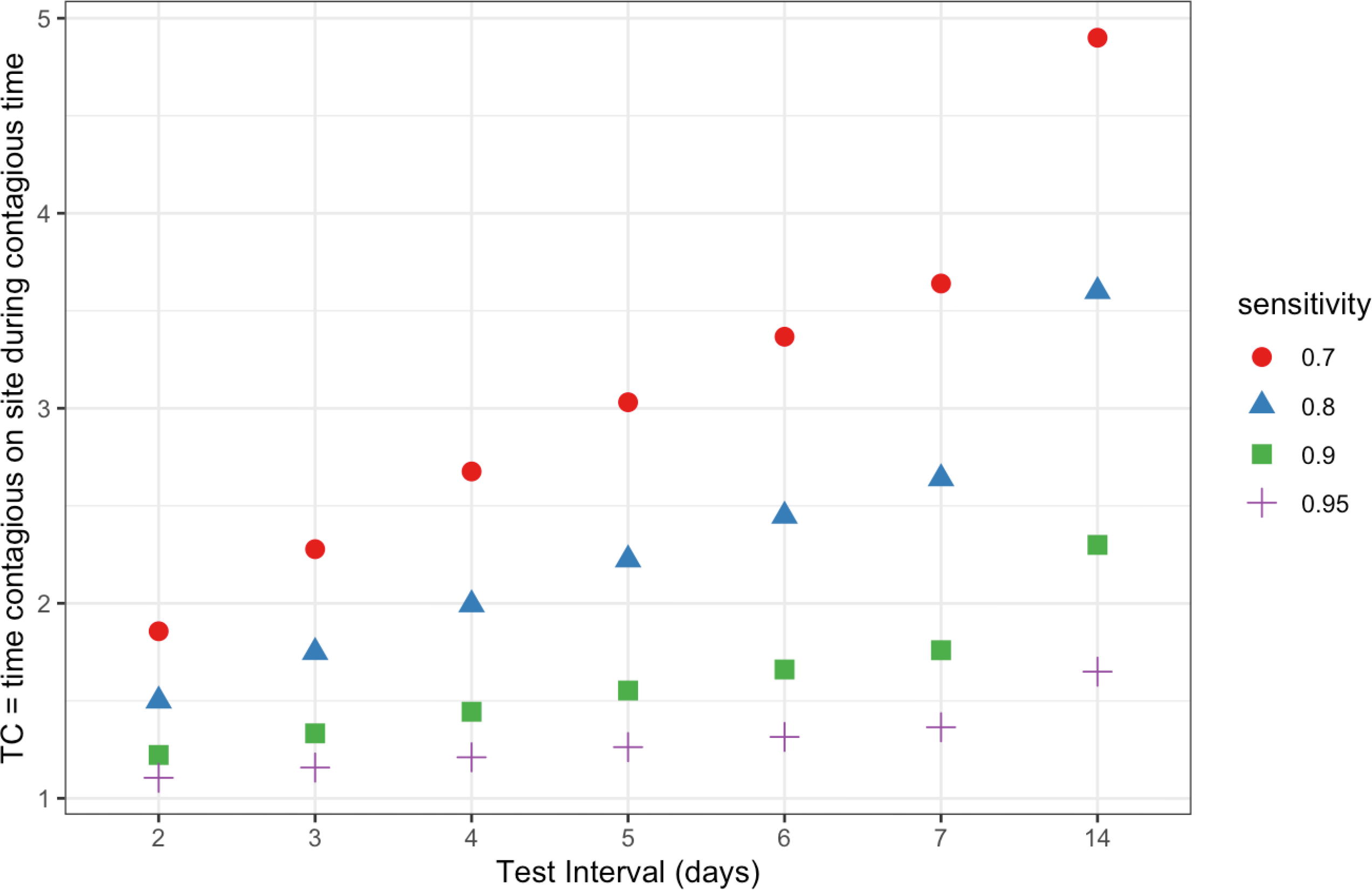
Expected time an infected employee remains on site. Based on a 1-day turnaround time and a contagious period of 14 days.

The expected number of days an employee is on site depends on sensitivity, testing interval, and turnaround time (Figure 7). If an infected contagious employee is on site and the testing interval is larger than the infectious period, then testing will not detect the infected employee. Here, we have modelled 14 and 21 days as that is the current estimated typical infectious period for SARS-CoV-2. For example, if on average, an infected individual is contagious for 14 days, the testing interval needs to be less than every 14 days in order to have a chance to identify the person. If the turnaround time to a test result is 1 day, and the test interval is 7 days (weekly) then the expected number of days an infected contagious employee remains on site is 2.6 days, for a test with 80% sensitivity, as opposed to 14 days with no testing during the infectious period (Supplementary Table 10). Thus, testing would reduce the time an infected employee was on site and contagious by 11.4 days (14− 2.6), an 81% (11.4/14) reduction of the time on site compared to no testing. If instead the test interval is every other day (2 days), then the expected number of days an infected contagious employee remains on site is 1.5 days (Supplementary Table 10). Testing has reduced the time on site and contagious 12.5 days (14− 1.5), an 89% (12.5/14) reduction.

Supplementary Table 11 shows the same modeling for a 21-day contagious period. Supplementary Figures 1 and 2 show a graphical representation of testing over the contagious period as well as the formula used for calculating the time a contagious person would be onsite.

### Management of screen positive employees

For employees that screen positive, initial management would be to instruct the employee to isolate (not return on site). The percent of screen positive employees will be low when prevalence is low. Importantly, the chance that the screen positive employee is infected is also low. The (post-test) probability that the employee is infected (true positive result) is shown as a function of pre-test prevalence for a range of sensitivity and specificity values in Figure 8. The expected test results and true status were determined across prevalence’s for a test sensitivity of 70% (Supplementary Table 4), 80% (Supplementary Table 5), 90% (Supplementary Table 6), and 95% (Supplementary Table 7).

**Figure 8.**
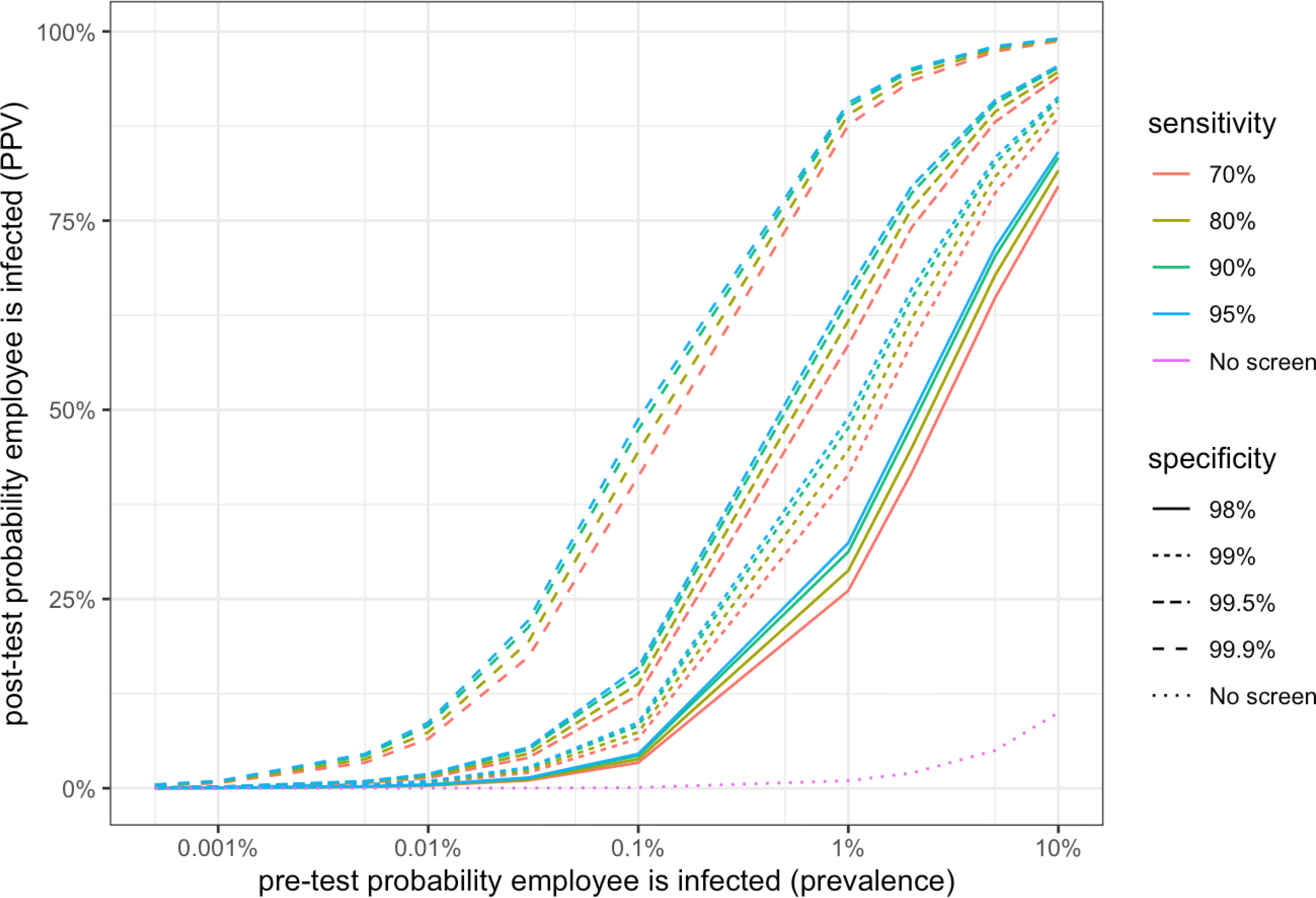
Probability that a single screen positive employee is infected with COVID-19 [true positive result] versus log10(prevalence) for a range of sensitivity and specificity values.

The implications for management of screen positives can be easier to understand when applying the modeled variables to a hypothetical population. Following is an example employer that had 1000 employees isolating before implementing an employee screening program to return to work. Implementation of a screening program with an 80% sensitivity and 99.5% specificity with a prevalence of 0.5% would result in ∼9 (4 TP + 5 FP) test positive employees (posttest) (Table 5). The impact of this can be viewed from both the employer’s and employee’s perspective. For the employer, instead of 1000 employees in isolation, they now have 991 employees that can return on site and only 9 employees self-isolating. For those 9 employees screening positive, 55.4% are predicted to be false positives, with a 44.6% chance that the employee is truly infected (Supplementary Table 5). In this case, this means that it is more likely that a screened positive employee is a false positive than a true positive. A positive screening result has implications for the employee in terms of triggering isolation and possibly confirmatory testing. Further, it could also impact any family, friends, and coworkers that have had recent contact with the screen-positive employee. Thus, it may be important for employers to message that employees who test positive have a reasonable chance that they are uninfected. Calculation details for this example are described in Supplementary Materials.

### Application of modelling to implement an employee screening program

As a final step, we applied the above modeling to a hypothetical population as an example of how to devise an example strategy for implement of SARS-CoV-2 screening and employee return to onsite work. Figure 9 gives an overview of a potential SARS-CoV-2 screening program for a return to work. To reduce the risk for onsite transmission while the community prevalence remains high, employees that can work effectively remotely may continue to do so until such time as the prevalence drops substantially or a vaccine becomes available. For employees returning on site, a baseline screen would be performed before returning on site by employees lacking COVID-19 symptoms and passing a thermal scan. Any employee that has COVID-19 symptoms, fails the thermal scan, or screens positive would self-isolate and follow Employer Health and Safety protocols for suspected COVID-19 infection. Employees that clear the baseline screening would return to onsite work and undergo regular SARS-CoV-2 screening.

**Figure 9.**
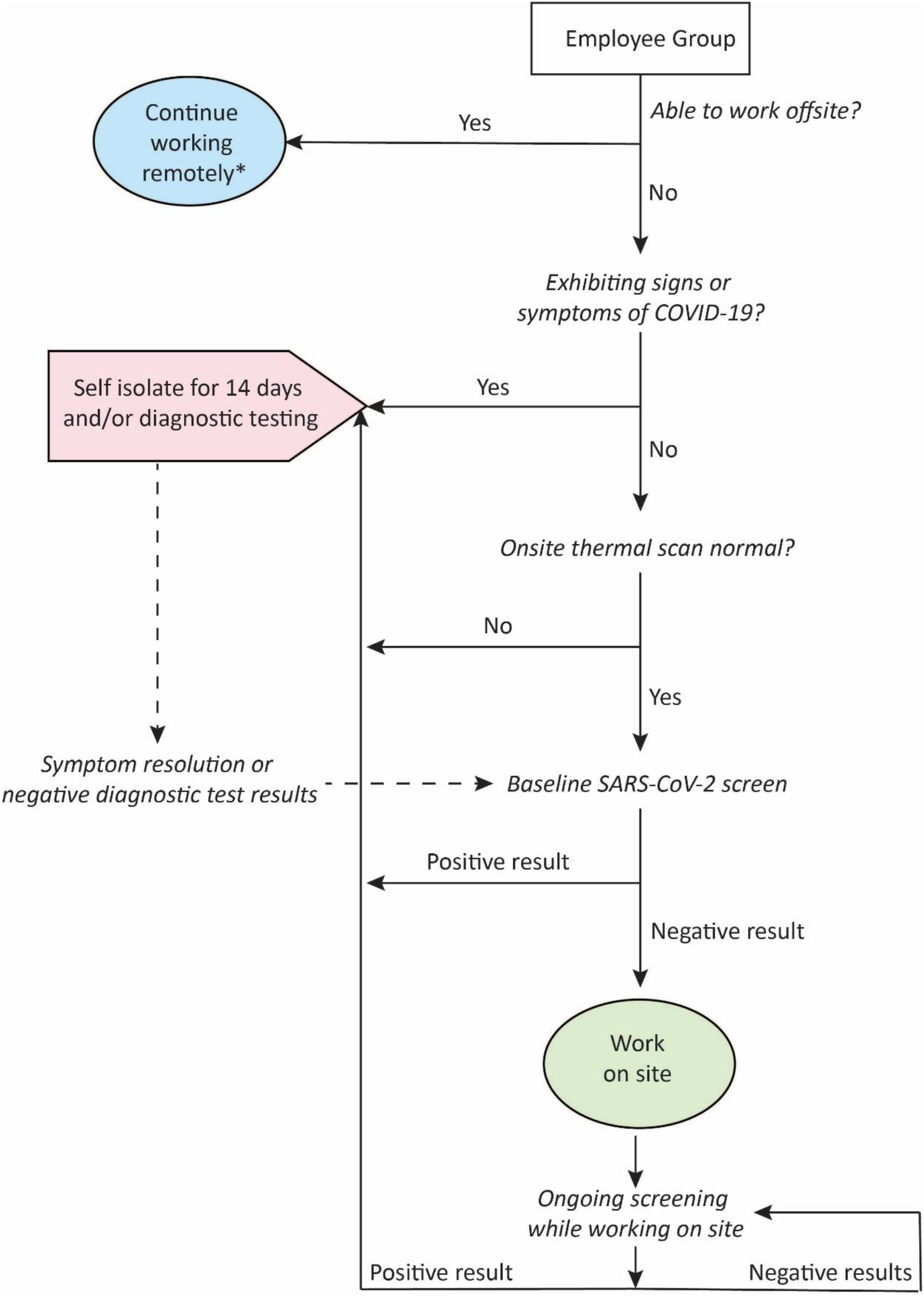
Return to work workflow incorporating baseline and repeat SARS-CoV-2 screening. * Until prevalence drops below set threshold or vaccine becomes available

In a simple example, an employer with 1000 individuals is interested in a return to onsite work. Their local community has a population of around 3 million, and recent testing information reports that around 600,000 tests were performed in the last two weeks with a 14-day rolling average percentage positive rate of 5%. Assuming these are unique individuals and that this reflects all current positive cases, this suggests a community prevalence close to 1%. However, there is likely a significant number of unrecognized asymptomatic cases in the community, so the true prevalence may be higher. The employer is planning on having employees displaying symptoms stay home, and thus, the prevalence among his asymptomatic employees could be lower than the community prevalence estimate. Based on this, the employer used a prevalence estimate of 1% but evaluated a prevalence range of 0.5% to 2% to account for potential variation in the true community prevalence. In the absence of an employee screening program, the probability of having at least one infected employee on site for a group of 1000 employees is 100% at a prevalence of 1.0% (99.3% at 0.5% prevalence and 100% at a prevalence of 2.0%; Supplementary Table 5). The employee determined that their threshold of acceptable risk for having at least one infected employee onsite was 10%. As such, bringing back 1000 employees in the absence of an employee screening program would have an intolerable level of risk (risk above the established threshold). One way to reduce risk is to reduce group size, however, the only scenario that would reduce risk below their 10% threshold would be a group size of 20 and assuming a prevalence of 0.5%. Thus, the employer next considered the benefit of an employee screening program.

For this example, SARS-CoV-2 testing is performed one time on a group of 1000 asymptomatic employees before returning to work on site. The test has a sensitivity of 80%, specificity of 99.5%, and a one-day TAT to get results. A baseline screen of the 1000 employees with a prevalence of 1.0% would result in 13 employees with a positive screen, and 987 with a negative screen (Supplementary Table 5). Those who are test positive are required to self-isolate and those who are test negative may return to work on site. The probability that at least one of the 987 screen negative employees that would return on site is infected is 86.5%. The workplace prevalence on Day 1 on site is reduced from 1.0% to 0.2% by baseline testing prior to returning on site, but there is still an 86.5% chance than an infected employee will come on site. If the employer reduced the group size return to onsite work to 50, assuming a 1.0% prevalence, the probability of at least one false negative (9.5%) would fall below their risk threshold; at 2.0% prevalence with 50 employees, the risk would be 18.2%, which is above their threshold. Thus, baseline screening has reduced the risk of an infected person returning on site, and if having a small group return is feasible, this approach may be acceptable. Figure 5 may be helpful in visualizing what group sizes can be used to keep the probability of at least one infected person on site below a set threshold within the prevalence range being considered.

If the employer would like to reduce the risk of an infected screen negative employee in a fixed group size returning, there are two main options, use a test with higher sensitivity or repeat testing before return. As shown in Table 4, repeat testing can help increase the probability an infected employee will be detected by each test cycle, thereby reducing the chance of an infected person screening negative and returning on site. Using a test with 80% sensitivity, after a second test, the probability of detecting an infected employee is 96%. For example, in a baseline screen of the 50 employees with a prevalence of 0.5%, the chance an infected screen negative employee returns on site is 4.9% (Supplementary Table 5) when test sensitivity is 80% versus less than 1.2% (Supplementary Table 7) is the test is performed twice in a row before return to work, resulting in a combined test sensitivity of 96% (>95%) (Table 4). This strategy would help reduce the probability of an infected person returning on site. Frequent testing on site reduces the time and infected person is on site (Supplementary Table 10).

To reduce the cost of frequent testing, sample pooling could be considered. For individual samples at a prevalence of 1.0%, there would be 13 positive screen results, 987 negative screen results, and an 86% risk for at least one infected person on site (Supplementary Table 8 and 9, grey shaded rows indicate individual samples). In comparison, with 1-stage sample pooling with 5 samples per pool, there would be 44 positive screen results, 956 negative screen results, and an 86% risk for at least one infected person on site (Supplementary Table 8). Thus, 1-stage pooling would reduce the number of employees returning to onsite work with no change in the probability that at least one of them is infected. With 2-stage sample pooling with 5 samples per pool, there would be 6 positive screen results, 994 negative screen results, and a 98% risk for at least one infected person on site (Supplementary Table 9). Thus, 2-stage pooling would increase the number of employees returning to onsite work but also increase the probability that at least one of them is infected compared with individual sample testing.

## DISCUSSION

In this paper, we describe statistical modeling we have performed to evaluate the benefit of introducing an employee SARS-CoV-2 screen to enable a return to onsite work. We evaluated a range of different variables, to address the limited information on SARS-CoV-2 transmissibility and prevalence and cover a range of variables involved in implementing a screening test. This modeling has been instrumental in developing the return to work policy for our employees.

The ability of a business to return to onsite work and their specific policy in the era of COVID-19 will depend on several factors. First, national and local governing bodies may have specific rules and regulations that businesses need to follow^38, 39^, which may preclude some businesses from returning to onsite work while the community prevalence remains above a specific threshold. For example, California tracks county-level data for several metrics related to positivity rates and hospitalizations^19^. Counties reporting numbers above set thresholds are placed on a “watch list” for monitoring worsening trends^18, 19^. Counties on the watch list have limitations on which indoor businesses can operate. The CDC has released interim guidance for businesses and employers responding to COVID-19^39^: Conducting daily health checks; conducting a hazard assessment of the workplace; encouraging employees to wear cloth face coverings in the workplace, if appropriate; implementing policies and practices for social distancing in the workplace; and improving the building ventilation system. The ability to implement social distancing and other precautionary measures may influence a given employer’s return to work policy, as well as the necessity for employees to be on site for business resumption. Some employers may be able to keep a proportion of their employees working remotely to reduce the required group size and the risk of workplace infection^40^. Finally, the desired degree of risk reduction or risk tolerance threshold will vary between employers. Employers for whom a single onsite infected employee may trigger a site shutdown and loss of business may desire to minimize risk to whatever degree is possible.

The strength of this study is in the variety of variables modeled and the range of values modeled. The findings here are consistent with a recent paper that modeled COVID-19 screening strategies that would allow a safe re-opening of US college campuses^15^. Multiple studies have reported test frequency has more impact on reducing COVID-19 infections than test sensitivity, and that rapid and frequent screening should be prioritized for general population screening^15, 23, 41^. Importantly, the optimal screening frequency is reliant upon successful adoption of behavioral interventions^15, 42^. Here, the intention is that other employers or groups could evaluate a scenario with a similar community prevalence and group size to one presented here to help inform their policy for returning on site. However, it is important to note that many epidemiological variables related to COVID-19 remain unknown or are based on limited information. While data suggests that the viral load may be similar in symptomatic and asymptomatic individuals with COVID-19,^43, 44^ there is limited data on the viral load over time in pre-symptomatic and asymptomatic individuals^44^. As test sensitivity is likely to be strongly linked to viral load, this supports that a screen in asymptomatic individuals may be able to achieve similar sensitivities to established diagnostic tests developed for the symptomatic population. But since this is not yet established and because many other variables remain uncertain, we recommend consideration across a range for each variable to account for this limitation.

When implementing a screening program, there are some important considerations and some potentials for harm that should be considered. With screening tests, there is typically a balance between sensitivity and specificity. A higher sensitivity (probability that an infected individual will have a positive test result) can generally be obtained by tolerating a lower specificity (probability that an uninfected individual will have a negative test result), and vice versa. There are implications of this for the employer and employees. Thus, while the focus of screening is to minimize the potential harm due to spread of COVID-19 in employees returning to work, it is also useful to consider the number of employees unnecessarily self-isolating. With high sensitivity, the chance of an infected employee returning on site is minimized. However, a greater proportion of the employee pool will screen positive, and most of these employees would be false positives and unnecessarily self-isolating. Similarly, more frequent testing would lead to more false positive results. High test specificity reduces the number of uninfected employees with a [false] positive test result that self-isolate unnecessarily. Further, a positive screening result may cause significant personal stress to employees. The harm of unnecessary self-isolation would impact the individual person and the employer but would not contribute to the risk of spread of COVID-19 in the workplace. Although baseline screening reduces the initial risk for an infected employee returning on site, there remains a potential for harm for employees returning on site. The potential for harm is that one pre-symptomatic or asymptomatic infected employee (with a false negative test result) returns on site and the virus is transmitted to other employees prior to the infected employee subsequently testing positive. The exact risk of transmission is unknown but could be minimized by workplace procedures such as social distancing, masks, and contact free doors. Another potential for harm is that an uninfected individual on site can become infected from the community. The number and time infectious employees are on site can be minimized though frequent testing and/or using a high sensitivity test.

A primary concern with implementation of a screening test, particularly one requiring frequent repeat tests, is the cost. The cost of a screening program needs to be balanced against the benefit in terms of risk reduction. Since resources and capacity for frequent testing may be limited, we explored the impact of sample pooling. Here, we evaluated both a 2-stage (Dorfman) and 1-stage approach to sample pooling. The Dorfman 2-stage pooled testing procedure will reduce the total number of tests but may have an increased chance of an infected employee on site compared with individual testing depending on prevalence. Despite this, it may be beneficial if it permits more frequent testing. It is also important to consider the logistics and feasibility of pooled sample testing. The Dorfman 2-stage procedure where follow-up testing is required may be more difficult from a logistics perspective. Importantly, current guidance from the CDC and FDA is that if sample pooling is used, when a test result for a pool is indeterminate or positive, all samples in the pool need to be retested individually (Dorfman 2-stage approach)^35, 36^.

In recognition of the fact that organizations may want to incorporate SARS-CoV-2 screening as part of a strategy to reduce workplace risk, the CDC has released guidance related to SARS-CoV-2 testing for non-healthcare workplaces^45^. In turn, the FDA released guidance on how to validate molecular tests that will be submitted for Emergency Use Authorization (EUA), including how to evaluate and validate sample pooling^35^. The FDA notes that for asymptomatic SARS-CoV-2 screening, a highly sensitive test is desirable but if this is not feasible, then serial testing on different days or with different tests should be considered^44^. The FDA encourages test developers to consider validating their tests intended for screening asymptomatic individuals^46^. The first COVID-19 test with EUA for the asymptomatic population was approved in July of 2020^47-49^. SARS-CoV-2 screens intended for use in the asymptomatic population will be a key part of broad screening initiatives.

In conclusion, we modeled of a range of key metrics that can be used to guide development of policy for a return to onsite work. We evaluated factors such as prevalence, group size, screening performance, screening frequency, and sample pooling on the risk of an infected person coming on site. We are using this modeling to develop an online resource to allow users to input their specific variables to enable them to evaluate potential testing schedules for a safe return to work. There are a multitude of variables and considerations for employers opting to return to onsite work, and we hope that the work presented here can be of use in the current COVID-19 pandemic.

## Data Availability

We are currently developing an online version of a tool so that people can explore the modeling presented here.

## ACKNOWLEDGEMENTS

The authors would like to thank Gerry Gray (Data-Fi, LLC) for his input on the modeling and graphs, Aida Yazdanparast for her work developing the online tool, Lucia Speroni for her assistance with reference identification and review of the manuscript, and Johan Surtihadi for critical review of the manuscript.

