## Supplementary Material for "A Working Model to Inform Risk-Based Back to Work Strategies"

### SUPPLEMENTARY METHODS

***Baseline SARS-CoV-2 testing, individual samples – Example of calculations***

Following is an example employer that had 1000 employees (NE) isolating before implementing an employee screening program to return to work. The pre-test prevalence of COVID-19 is 0.5%. The employer implements a screening program testing for SARS-CoV-2 with test sensitivity and specificity of 80% and 99.5%. The number of infected and “healthy” (uninfected) employees is unknown but one can calculate the expected number for each based on estimated prevalence, 0.5% (0.005). The expected number of infected versus healthy employees is calculated as:

- Number infected (NI) = NE×prev = 1000×0.005 = 5
- Number healthy (NH) = NE×(1–prev) = 1000×0.995 = 995

Most employees are expected to be healthy since the prevalence is very small. If the 1000 employees are tested using a test with sensitivity = 80% (0.8) and specificity =99.5% (0.995) then the expected number of TP, FP, FN, and TN results (Table 5) are calculated using the formulas from Supplementary Table 1 and presented using the format of Supplementary Table 2.

- TP = number infected × sens = 5×0.80 = 4
- FN = number infected × (1–sens) = 5×0.20 = 1
- TN = number healthy × spec = 995×0.995 = 990
- FP = number healthy × (1–spec) = 995×0.005 = 5

In practice, the employer will not know who is infected versus healthy so they will not know the number of TP, FP, FN, and TN. They will know the number of test positives versus test negatives employees. When prevalence is very low, the number and percent of test positives will be very low and the number and percent of test negatives will be high, calculated as:

- npos=number of test positives = TP+FP = 4+5 = 9
    - Percent of test positives = Number of test positives/number of employees = 9/1000 or 0.9%
- nneg=number of test negatives = FN+TN = 1+990 = 991
    - Percent of test positives = Number of test positives/number of employees = 991/1000 or 99.1%

It is important to note that the test positive rate (npos/NE) is not the same as prevalence (NI/NE). The test positive rate is lower than prevalence when prevalence is very low and higher than prevalence when prevalence is higher.

While the employer and employee will not know who is infected, knowing the test results informs the employee about the chance they are infected. The pretest probability that an employee is infected is 0.5% (prevalence). The posttest chance they are infected depends on the test result. The chance that a test negative employee is infected (prevalence in test negatives) and the chance that a test positive employee is infected (prevalence in test positives) are calculated using the formulas below. If an employee is test negative, there is a small chance (0.1%) they are infected; if they are test positive, there is a 44.6% chance they are infected.

- chance that a test negative employee is infected (1–NPV)

- 1− (1−prev)×spec/[(1−prev) ×spec + prev× (1−sens)]

  = 1− (0.995×0.995)/(0.995×0.995 + 0.005×0.2) = 0.001 or 0.1%

- chance that a test positive employee is infected (PPV)
  - prev×sens/[prev×sens + (1−prev)×(1−spec)]

    = (0.005×0.8)/(0.005×0.8 + 0.995×0.005) = 0.446 or 44.6%

Often literature refers to false negative or false positive *rates*. It is important to clarify the group (denominator) for which this rate is calculated. In the example above, the number of false positive results is 5. The false positive rate can be calculated among different groups, for example

- among all employees: 0.5% (5/1000)
- among healthy employees (1−spec): 0.5% (5/995)
- among test positive employees (1−PPV): 56% (5/9)

Similarly, the false negative "rate" can be calculated among different groups, for example

- among all employees: 0.1% (1/1000)
- among infected employees (1−sens): 20% (1/5)
- among test negative employees (NPV): 0.1% (1/991)

***Baseline SARS-CoV-2 testing - pooled samples***

The risk of having any infected employee on site using the 1-stage and 2-stage pooled sample approach is calculated by simulation assuming individual outcomes are independent Bernoulli trials. Ten thousand simulated populations of size NE were generated. For each simulated population, the number of FN, FP, TN and TP results across individual declared results were calculated along with an indicator variable if FN>0. The reported expected number of results in Supplementary Tables 8 and 9 were calculated as average values across all simulated populations.

In each single simulated population of size NE, an individual true status and an individual declared test result based on the pooled testing scheme (1-stage and 2-stage) was generated. An individual test result is called 'declared' because it may be an assigned test result based on a pooled test result as opposed to an outcome from an individual level test. First, the individual true status is assigned as infected with probability P. Next, the NE simulated individuals are assigned to a pool of size k. There will be NE/k sample pools. True pool status is assigned as infected if any [at least one] simulated individual in the pool has a true individual status of infected. For pools assigned healthy, the pooled test result is assigned as negative with probability pool specificity (sp.k); for pools assigned infected, the pooled test result is assigned as positive with probability pool sensitivity (se.k). The stage 1 individual test result is assigned the pooled test result. The stage 2 individual test result is assigned the pooled test result when the pooled test result is negative. If the pooled test result is positive, the stage 2 individual test result is assigned based on the individual true status: if the individual true status is infected then the stage 2 individual test result is assigned positive with probability [individual] sensitivity (se); if the individual true individual status is healthy then the stage 2 individual test result is assigned negative with probability [individual] specificity (sp).

Summary results from the simulation are reported in Supplementary Tables 8 and 9. In some simulated populations when prevalence is low, there were no declared positive individual result generated. Therefore, the average number of positive declared individual results and FP declared individual results were calculated conditional on at least one positive declared individual result in the simulated population. The percent of time there was no declared positive individual result generated across all simulated populations is reported in the last column of each table.

**Supplementary Table 1. Formulas**

| No. | Formula Name | Formula Description/Notes | Formula |
|---|---|---|---|
| 1 | No Testing – Probability of at least one infected | Calculated based on modeling the number of infected individuals out of NE as following a binomial distribution with parameters NE and P. Z=number infected out of NE. | $\Pr(Z\geq1) = 1 - [\Pr(Z=0)]$<br>$= 1 - (1-P)^{NE}$ |
| 2 | Cross tabulation expected test results vs true status | Expected counts are calculated based on modeling the number of test positive employees out of NI infected individuals as following a binomial distribution with parameters NI and sensitivity. and the number of test negative employees out of NH uninfected individuals as following a binomial distribution with parameters NH and specificity. P, se, and sp are represented as decimals between 0 and 1. | $NI = NE \times P$<br>$NH = NE \times (1- P)$<br>$TP = NI \times se = NE \times P \times se$<br>$FN = NI \times (1- se) = NE \times P \times (1- se)$<br>$FP = NH \times (1- sp) = NE \times (1- P) \times (1- sp)$<br>$TN = NH \times sp = NE \times (1- P) \times sp$<br>$Npos = TP + FP = NE \times P \times se + NE \times (1- P) \times (1- sp)$<br>$Nneg = FN + TN = NE \times P \times (1- se) + NE \times (1- P) \times sp$ |
| 3 | Negative predictive value (NPV) | Likelihood of a screen negative employee being uninfected. P, se, and sp are represented as decimals between 0 and 1. | $NPV=(1-P)\times sp/[(1-P)\times sp + P\times(1-se)]$ |
| | | Likelihood of a screen negative employee being infected. | $1-NPV$ |
| 4 | Positive predictive value (PPV) | Likelihood of a screen positive employee being infected. P, se, and sp are represented as decimals between 0 and 1. | $PPV=P\times se/[P\times se + (1-P)\times(1-sp)]$ |
| | | Likelihood of a screen positive employee being uninfected. | $1-PPV$ |
| 5 | Baseline screen, individual samples – Probability of at least one infected | The risk of having any infected employee on site is the probability of having at least 1 false negative employee out of Nneg test negative employees on site. Calculated by modeling the number of infected [FN] individuals out of Nneg test negative | $\Pr(Z\geq1) = 1 - [\Pr(Z=0)]$<br>$= 1 - [1 - (1 - NPV)^{Nneg}]$<br>$= 1 - NPV^{Nneg}$ |

| No. | Formula Name | Formula Description/Notes | Formula |
|---|---|---|---|
| | | employees as following a binomial distribution with parameters Nneg and 1–NPV, where Nneg is treated as a known quantity. Z=number infected out of Nneg. | |
| 6 | Sample pooling - sensitivity | Test sensitivity for pooled sample was estimated as sensitivity for individual samples minus 1.5% and 2.5% for k=5 and 10, respectively. Se is represented as a decimal between 0 and 1. | se.5 = se – 0.015<br><br>se.10 = se – 0.025 |
| 7 | Sample pooling – prevalence of infected pools | The prevalence of infected pools is the pre-test probability that a pool is infected. A pool is infected if at least 1 sample in the pool is from an infected employee. The probability that a pool is infected is calculated by modeling the number of infected samples out of k samples in the pool as following a binomial distribution with parameters k and individual pre-test prevalence P. Z=number infected samples out of k. | $Pr(Z \geq 1) = 1 - [Pr(Z=0)] = 1 - (1 - P)^k$ |
| 8 | Expected number of days an infected employee on site | The expected number of days an infected employee will be on site before being detected (TC) depends on se, the test interval (TI) and turnaround time (TA). m is the number of tests per infected and contagious time | $TC = (TA)\,se + (TA+TI)(1-se)se + (TA+2TI)(1-se)^2 se + \cdots + (TA+(m-1)TI)(1-se)^{m-1} se + TINF(1-se)^m$<br><br>m = 1 + integer part of ((TINF - 1)/TI) |

**Supplementary Table 2. Cross tabulation of expected SARS-CoV-2 Test Result versus the true status**

| | | **True Status** | | **Total** |
|---|---|---|---|---|
| | | **Infected** | **Uninfected "Healthy"** | |
| **Test Result** | **Test Positive** | TP | FP | Npos |
| | **Test Negative** | FN | TN | Nneg |
| | **Total** | NI | NH | NE |

Based on employees tested (NE), prevalence (P), test sensitivity, and test specificity. FN, false negative; FP, false positive; NH, number of "healthy" employees, where healthy means uninfected with respect to SARS-CoV-2; NI, number of infected employees out of NE screened at point in time; Npos, number of test positive employees; Nneg, number of test negative employees; TN, true negative; TP, true positive.

**Supplementary Table 3. Probability an infected employee on site will be detected by each test on site**

| | **Probability calculation** |
|---|---|
| **By Test 1** | se |
| **By Test 2** | se + se(1-se) |
| **By Test 3** | se + se(1-se) + se(1-se)$^2$ |
| **By Test 4** | se + se(1-se) + se(1-se)$^2$ + se(1-se)$^3$ |
| … | |
| **By Test m** | se + se(1-se) + se(1-se)$^2$ + ⋯ + se(1-se)$^{m-1}$ |

m, number of times individual tested per time infected; se, sensitivity of test.

**Supplementary Table 4. Risk comparison between no screening and baseline screen with a 70% sensitivity and 99.5% specificity**

| | | No Screen | | Baseline Screening Test: 70% sensitivity, 99.5% specificity | | | | | | |
|---|---|---|---|---|---|---|---|---|---|---|
| | | | | Screen negatives out of NE "return to work" | | | | Screen positives out of NE "isolate/stay home" | | |
| Pre-test Prevalence | Group Size NE | Number COVID19 Infected | Prob. at least 1 infected | Prevalence in screen negatives, 1 – NPV | Number TN | Number FN | Prob. at least 1 FN | Prevalence in screen positives, PPV | Number TP | Number FP |
| 0.001% | 20 | 0.0 | 0.02% | 0.000% | 19.9 | 0.0 | 0.0% | 0.1% | 0.0 | 0.1 |
| | 50 | 0.0 | 0.05% | 0.000% | 49.7 | 0.0 | 0.0% | 0.1% | 0.0 | 0.2 |
| | 100 | 0.0 | 0.10% | 0.000% | 99.5 | 0.0 | 0.0% | 0.1% | 0.0 | 0.5 |
| | 250 | 0.0 | 0.25% | 0.000% | 248.7 | 0.0 | 0.1% | 0.1% | 0.0 | 1.2 |
| | 500 | 0.0 | 0.50% | 0.000% | 497.5 | 0.0 | 0.1% | 0.1% | 0.0 | 2.5 |
| | 1000 | 0.0 | 1.00% | 0.000% | 995.0 | 0.0 | 0.3% | 0.1% | 0.0 | 5.0 |
| 0.010% | 20 | 0.0 | 0.20% | 0.003% | 19.9 | 0.0 | 0.1% | 1.4% | 0.0 | 0.1 |
| | 50 | 0.0 | 0.50% | 0.003% | 49.7 | 0.0 | 0.1% | 1.4% | 0.0 | 0.2 |
| | 100 | 0.0 | 1.00% | 0.003% | 99.5 | 0.0 | 0.3% | 1.4% | 0.0 | 0.5 |
| | 250 | 0.0 | 2.47% | 0.003% | 248.7 | 0.0 | 0.7% | 1.4% | 0.0 | 1.2 |
| | 500 | 0.0 | 4.88% | 0.003% | 497.5 | 0.0 | 1.5% | 1.4% | 0.0 | 2.5 |
| | 1000 | 0.1 | 9.52% | 0.003% | 994.9 | 0.0 | 3.0% | 1.4% | 0.1 | 5.0 |
| 0.100% | 20 | 0.0 | 1.98% | 0.030% | 19.9 | 0.0 | 0.6% | 12.3% | 0.0 | 0.1 |
| | 50 | 0.0 | 4.88% | 0.030% | 49.7 | 0.0 | 1.5% | 12.3% | 0.0 | 0.2 |
| | 100 | 0.1 | 9.52% | 0.030% | 99.4 | 0.0 | 3.0% | 12.3% | 0.1 | 0.5 |
| | 250 | 0.2 | 22.13% | 0.030% | 248.5 | 0.1 | 7.2% | 12.3% | 0.2 | 1.2 |
| | 500 | 0.5 | 39.36% | 0.030% | 497.0 | 0.2 | 13.9% | 12.3% | 0.4 | 2.5 |
| | 1000 | 1.0 | 63.23% | 0.030% | 994.0 | 0.3 | 25.9% | 12.3% | 0.7 | 5.0 |

| | | No Screen | | Baseline Screening Test: 70% sensitivity, 99.5% specificity | | | | | | |
|---|---|---|---|---|---|---|---|---|---|---|
| | | | | Screen negatives out of NE “return to work” | | | | Screen positives out of NE “isolate/stay home” | | |
| **Pre-test Prevalence** | **Group Size NE** | **Number COVID19 Infected** | **Prob. at least 1 infected** | **Prevalence in screen negatives, 1 – NPV** | **Number TN** | **Number FN** | **Prob. at least 1 FN** | **Prevalence in screen positives, PPV** | **Number TP** | **Number FP** |
| 0.500% | 20 | 0.1 | 9.54% | 0.151% | 19.8 | 0.0 | 3.0% | 41.3% | 0.1 | 0.1 |
| | 50 | 0.2 | 22.17% | 0.151% | 49.5 | 0.1 | 7.2% | 41.3% | 0.2 | 0.2 |
| | 100 | 0.5 | 39.42% | 0.151% | 99.0 | 0.2 | 13.9% | 41.3% | 0.4 | 0.5 |
| | 250 | 1.2 | 71.44% | 0.151% | 247.5 | 0.4 | 31.3% | 41.3% | 0.9 | 1.2 |
| | 500 | 2.5 | 91.84% | 0.151% | 495.0 | 0.8 | 52.8% | 41.3% | 1.8 | 2.5 |
| | 1000 | 5.0 | 99.33% | 0.151% | 990.0 | 1.5 | 77.7% | 41.3% | 3.5 | 5.0 |
| 1.000% | 20 | 0.2 | 18.21% | 0.304% | 19.7 | 0.1 | 5.8% | 58.6% | 0.1 | 0.1 |
| | 50 | 0.5 | 39.50% | 0.304% | 49.3 | 0.2 | 14.0% | 58.6% | 0.4 | 0.2 |
| | 100 | 1.0 | 63.40% | 0.304% | 98.5 | 0.3 | 26.0% | 58.6% | 0.7 | 0.5 |
| | 250 | 2.5 | 91.89% | 0.304% | 246.3 | 0.8 | 52.8% | 58.6% | 1.8 | 1.2 |
| | 500 | 5.0 | 99.34% | 0.304% | 492.5 | 1.5 | 77.7% | 58.6% | 3.5 | 2.5 |
| | 1000 | 10.0 | 100.00% | 0.304% | 985.0 | 3.0 | 95.0% | 58.6% | 7.0 | 5.0 |
| 2.000% | 20 | 0.4 | 33.24% | 0.612% | 19.5 | 0.1 | 11.3% | 74.1% | 0.3 | 0.1 |
| | 50 | 1.0 | 63.58% | 0.612% | 48.8 | 0.3 | 26.0% | 74.1% | 0.7 | 0.2 |
| | 100 | 2.0 | 86.74% | 0.612% | 97.5 | 0.6 | 45.2% | 74.1% | 1.4 | 0.5 |
| | 250 | 5.0 | 99.36% | 0.612% | 243.8 | 1.5 | 77.8% | 74.1% | 3.5 | 1.2 |
| | 500 | 10.0 | 100.00% | 0.612% | 487.6 | 3.0 | 95.1% | 74.1% | 7.0 | 2.5 |
| | 1000 | 20.0 | 100.00% | 0.612% | 975.1 | 6.0 | 99.8% | 74.1% | 14.0 | 4.9 |

FN, false negative; FP, false positive; NPV, negative predictive value; PPV, positive predictive value; prob, probability; TN, true negative; TP, true positive.

Numbers in the table represent expected values, or what would be observed on average.  Actual numbers will vary.

**Supplementary Table 5. Risk comparison between no screening and baseline screen with an 80% sensitivity and 99.5% specificity**

| | | No Screen | | Baseline Screening Test: 80% sensitivity, 99.5% specificity | | | | | | |
|---|---|---|---|---|---|---|---|---|---|---|
| | | | | Screen negatives out of NE "return to work" | | | | Screen positives out of NE "isolate/stay home" | | |
| Pre-test Prevalence | Group Size NE | Number COVID19 Infected | Prob. at least 1 infected | Prevalence in screen negatives, 1 – NPV | Number TN | Number FN | Prob. at least 1 FN | Prevalence in screen positives, PPV | Number TP | Number FP |
| 0.001% | 20 | 0.0 | 0.02% | 0.000% | 19.9 | 0.0 | 0.0% | 0.2% | 0.0 | 0.1 |
| | 50 | 0.0 | 0.05% | 0.000% | 49.7 | 0.0 | 0.0% | 0.2% | 0.0 | 0.2 |
| | 100 | 0.0 | 0.10% | 0.000% | 99.5 | 0.0 | 0.0% | 0.2% | 0.0 | 0.5 |
| | 250 | 0.0 | 0.25% | 0.000% | 248.7 | 0.0 | 0.0% | 0.2% | 0.0 | 1.2 |
| | 500 | 0.0 | 0.50% | 0.000% | 497.5 | 0.0 | 0.1% | 0.2% | 0.0 | 2.5 |
| | 1000 | 0.0 | 1.00% | 0.000% | 995.0 | 0.0 | 0.2% | 0.2% | 0.0 | 5.0 |
| 0.010% | 20 | 0.0 | 0.20% | 0.002% | 19.9 | 0.0 | 0.0% | 1.6% | 0.0 | 0.1 |
| | 50 | 0.0 | 0.50% | 0.002% | 49.7 | 0.0 | 0.1% | 1.6% | 0.0 | 0.2 |
| | 100 | 0.0 | 1.00% | 0.002% | 99.5 | 0.0 | 0.2% | 1.6% | 0.0 | 0.5 |
| | 250 | 0.0 | 2.47% | 0.002% | 248.7 | 0.0 | 0.5% | 1.6% | 0.0 | 1.2 |
| | 500 | 0.0 | 4.88% | 0.002% | 497.5 | 0.0 | 1.0% | 1.6% | 0.0 | 2.5 |
| | 1000 | 0.1 | 9.52% | 0.002% | 994.9 | 0.0 | 2.0% | 1.6% | 0.1 | 5.0 |
| 0.100% | 20 | 0.0 | 1.98% | 0.020% | 19.9 | 0.0 | 0.4% | 13.8% | 0.0 | 0.1 |
| | 50 | 0.0 | 4.88% | 0.020% | 49.7 | 0.0 | 1.0% | 13.8% | 0.0 | 0.2 |
| | 100 | 0.1 | 9.52% | 0.020% | 99.4 | 0.0 | 2.0% | 13.8% | 0.1 | 0.5 |
| | 250 | 0.2 | 22.13% | 0.020% | 248.5 | 0.0 | 4.9% | 13.8% | 0.2 | 1.2 |
| | 500 | 0.5 | 39.36% | 0.020% | 497.0 | 0.1 | 9.5% | 13.8% | 0.4 | 2.5 |
| | 1000 | 1.0 | 63.23% | 0.020% | 994.0 | 0.2 | 18.1% | 13.8% | 0.8 | 5.0 |

| | | No Screen | | Baseline Screening Test: 80% sensitivity, 99.5% specificity | | | | | | |
|---|---|---|---|---|---|---|---|---|---|---|
| | | | | Screen negatives out of NE “return to work” | | | | Screen positives out of NE “isolate/stay home” | | |
| Pre-test Prevalence | Group Size NE | Number COVID19 Infected | Prob. at least 1 infected | Prevalence in screen negatives, 1 – NPV | Number TN | Number FN | Prob. at least 1 FN | Prevalence in screen positives, PPV | Number TP | Number FP |
| 0.500% | 20 | 0.1 | 9.54% | 0.101% | 19.8 | 0.0 | 2.0% | 44.6% | 0.1 | 0.1 |
| | 50 | 0.2 | 22.17% | 0.101% | 49.5 | 0.0 | 4.9% | 44.6% | 0.2 | 0.2 |
| | 100 | 0.5 | 39.42% | 0.101% | 99.0 | 0.1 | 9.5% | 44.6% | 0.4 | 0.5 |
| | 250 | 1.2 | 71.44% | 0.101% | 247.5 | 0.2 | 22.1% | 44.6% | 1.0 | 1.2 |
| | 500 | 2.5 | 91.84% | 0.101% | 495.0 | 0.5 | 39.4% | 44.6% | 2.0 | 2.5 |
| | 1000 | 5.0 | 99.33% | 0.101% | 990.0 | 1.0 | 63.2% | 44.6% | 4.0 | 5.0 |
| 1.000% | 20 | 0.2 | 18.21% | 0.203% | 19.7 | 0.0 | 3.9% | 61.8% | 0.2 | 0.1 |
| | 50 | 0.5 | 39.50% | 0.203% | 49.3 | 0.1 | 9.5% | 61.8% | 0.4 | 0.2 |
| | 100 | 1.0 | 63.40% | 0.203% | 98.5 | 0.2 | 18.1% | 61.8% | 0.8 | 0.5 |
| | 250 | 2.5 | 91.89% | 0.203% | 246.3 | 0.5 | 39.4% | 61.8% | 2.0 | 1.2 |
| | 500 | 5.0 | 99.34% | 0.203% | 492.5 | 1.0 | 63.2% | 61.8% | 4.0 | 2.5 |
| | 1000 | 10.0 | 100.00% | 0.203% | 985.0 | 2.0 | 86.5% | 61.8% | 8.0 | 5.0 |
| 2.000% | 20 | 0.4 | 33.24% | 0.409% | 19.5 | 0.1 | 7.7% | 76.6% | 0.3 | 0.1 |
| | 50 | 1.0 | 63.58% | 0.409% | 48.8 | 0.2 | 18.2% | 76.6% | 0.8 | 0.2 |
| | 100 | 2.0 | 86.74% | 0.409% | 97.5 | 0.4 | 33.0% | 76.6% | 1.6 | 0.5 |
| | 250 | 5.0 | 99.36% | 0.409% | 243.8 | 1.0 | 63.3% | 76.6% | 4.0 | 1.2 |
| | 500 | 10.0 | 100.00% | 0.409% | 487.6 | 2.0 | 86.5% | 76.6% | 8.0 | 2.5 |
| | 1000 | 20.0 | 100.00% | 0.409% | 975.1 | 4.0 | 98.2% | 76.6% | 16.0 | 4.9 |

FN, false negative; FP, false positive; NPV, negative predictive value; PPV, positive predictive value; prob, probability; TN, true negative; TP, true positive.

Numbers in the table represent expected values, or what would be observed on average. Actual numbers will vary.

**Supplementary Table 6. Risk comparison between no screening and baseline screen with a 90% sensitivity and 99.5% specificity**

| | | No Screen | | Baseline Screening Test: 90% sensitivity, 99.5% specificity | | | | | | |
|---|---|---|---|---|---|---|---|---|---|---|
| | | | | Screen negatives out of NE "return to work" | | | | Screen positives out of NE "isolate/stay home" | | |
| Pre-test Prevalence | Group Size NE | Number COVID19 Infected | Prob. at least 1 infected | Prevalence in screen negatives, 1 – NPV | Number TN | Number FN | Prob. at least 1 FN | Prevalence in screen positives, PPV | Number TP | Number FP |
| 0.001% | 20 | 0.0 | 0.02% | 0.000% | 19.9 | 0.0 | 0.0% | 0.2% | 0.0 | 0.1 |
| | 50 | 0.0 | 0.05% | 0.000% | 49.7 | 0.0 | 0.0% | 0.2% | 0.0 | 0.2 |
| | 100 | 0.0 | 0.10% | 0.000% | 99.5 | 0.0 | 0.0% | 0.2% | 0.0 | 0.5 |
| | 250 | 0.0 | 0.25% | 0.000% | 248.7 | 0.0 | 0.0% | 0.2% | 0.0 | 1.2 |
| | 500 | 0.0 | 0.50% | 0.000% | 497.5 | 0.0 | 0.0% | 0.2% | 0.0 | 2.5 |
| | 1000 | 0.0 | 1.00% | 0.000% | 995.0 | 0.0 | 0.1% | 0.2% | 0.0 | 5.0 |
| 0.010% | 20 | 0.0 | 0.20% | 0.001% | 19.9 | 0.0 | 0.0% | 1.8% | 0.0 | 0.1 |
| | 50 | 0.0 | 0.50% | 0.001% | 49.7 | 0.0 | 0.0% | 1.8% | 0.0 | 0.2 |
| | 100 | 0.0 | 1.00% | 0.001% | 99.5 | 0.0 | 0.1% | 1.8% | 0.0 | 0.5 |
| | 250 | 0.0 | 2.47% | 0.001% | 248.7 | 0.0 | 0.2% | 1.8% | 0.0 | 1.2 |
| | 500 | 0.0 | 4.88% | 0.001% | 497.5 | 0.0 | 0.5% | 1.8% | 0.0 | 2.5 |
| | 1000 | 0.1 | 9.52% | 0.001% | 994.9 | 0.0 | 1.0% | 1.8% | 0.1 | 5.0 |
| 0.100% | 20 | 0.0 | 1.98% | 0.010% | 19.9 | 0.0 | 0.2% | 15.3% | 0.0 | 0.1 |
| | 50 | 0.0 | 4.88% | 0.010% | 49.7 | 0.0 | 0.5% | 15.3% | 0.0 | 0.2 |
| | 100 | 0.1 | 9.52% | 0.010% | 99.4 | 0.0 | 1.0% | 15.3% | 0.1 | 0.5 |
| | 250 | 0.2 | 22.13% | 0.010% | 248.5 | 0.0 | 2.5% | 15.3% | 0.2 | 1.2 |
| | 500 | 0.5 | 39.36% | 0.010% | 497.0 | 0.0 | 4.9% | 15.3% | 0.4 | 2.5 |
| | 1000 | 1.0 | 63.23% | 0.010% | 994.0 | 0.1 | 9.5% | 15.3% | 0.9 | 5.0 |

| | | No Screen | | Baseline Screening Test: 90% sensitivity, 99.5% specificity | | | | | | |
|---|---|---|---|---|---|---|---|---|---|---|
| | | | | Screen negatives out of NE “return to work” | | | | Screen positives out of NE “isolate/stay home” | | |
| **Pre-test Prevalence** | **Group Size NE** | **Number COVID19 Infected** | **Prob. at least 1 infected** | **Prevalence in screen negatives, 1 – NPV** | **Number TN** | **Number FN** | **Prob. at least 1 FN** | **Prevalence in screen positives, PPV** | **Number TP** | **Number FP** |
| 0.500% | 20 | 0.1 | 9.54% | 0.050% | 19.8 | 0.0 | 1.0% | 47.5% | 0.1 | 0.1 |
| | 50 | 0.2 | 22.17% | 0.050% | 49.5 | 0.0 | 2.5% | 47.5% | 0.2 | 0.2 |
| | 100 | 0.5 | 39.42% | 0.050% | 99.0 | 0.0 | 4.9% | 47.5% | 0.4 | 0.5 |
| | 250 | 1.2 | 71.44% | 0.050% | 247.5 | 0.1 | 11.8% | 47.5% | 1.1 | 1.2 |
| | 500 | 2.5 | 91.84% | 0.050% | 495.0 | 0.2 | 22.1% | 47.5% | 2.2 | 2.5 |
| | 1000 | 5.0 | 99.33% | 0.050% | 990.0 | 0.5 | 39.4% | 47.5% | 4.5 | 5.0 |
| 1.000% | 20 | 0.2 | 18.21% | 0.101% | 19.7 | 0.0 | 2.0% | 64.5% | 0.2 | 0.1 |
| | 50 | 0.5 | 39.50% | 0.101% | 49.3 | 0.0 | 4.9% | 64.5% | 0.4 | 0.2 |
| | 100 | 1.0 | 63.40% | 0.101% | 98.5 | 0.1 | 9.5% | 64.5% | 0.9 | 0.5 |
| | 250 | 2.5 | 91.89% | 0.101% | 246.3 | 0.2 | 22.1% | 64.5% | 2.2 | 1.2 |
| | 500 | 5.0 | 99.34% | 0.101% | 492.5 | 0.5 | 39.4% | 64.5% | 4.5 | 2.5 |
| | 1000 | 10.0 | 100.00% | 0.101% | 985.0 | 1.0 | 63.2% | 64.5% | 9.0 | 5.0 |
| 2.000% | 20 | 0.4 | 33.24% | 0.205% | 19.5 | 0.0 | 3.9% | 78.6% | 0.4 | 0.1 |
| | 50 | 1.0 | 63.58% | 0.205% | 48.8 | 0.1 | 9.5% | 78.6% | 0.9 | 0.2 |
| | 100 | 2.0 | 86.74% | 0.205% | 97.5 | 0.2 | 18.1% | 78.6% | 1.8 | 0.5 |
| | 250 | 5.0 | 99.36% | 0.205% | 243.8 | 0.5 | 39.4% | 78.6% | 4.5 | 1.2 |
| | 500 | 10.0 | 100.00% | 0.205% | 487.6 | 1.0 | 63.3% | 78.6% | 9.0 | 2.5 |
| | 1000 | 20.0 | 100.00% | 0.205% | 975.1 | 2.0 | 86.5% | 78.6% | 18.0 | 4.9 |

FN, false negative; FP, false positive; NPV, negative predictive value; PPV, positive predictive value; prob, probability; TN, true negative; TP, true positive.

Numbers in the table represent expected values, or what would be observed on average. Actual numbers will vary.

**Supplementary Table 7. Risk comparison between no screening and baseline screen with 95% sensitivity and 99.5% specificity**

| | | No Screen | | Baseline Screening Test: 95% sensitivity, 99.5% specificity | | | | | | |
|---|---|---|---|---|---|---|---|---|---|---|
| | | | | Screen negatives out of NE "return to work" | | | | Screen positives out of NE "isolate/stay home" | | |
| Pre-test Prevalence | Group Size NE | Number COVID19 Infected | Prob. at least 1 infected | Prevalence in screen negatives, 1 – NPV | Number TN | Number FN | Prob. at least 1 FN | Prevalence in screen positives, PPV | Number TP | Number FP |
| 0.001% | 20 | 0.0 | 0.02% | 0.000% | 19.9 | 0.0 | 0.0% | 0.2% | 0.0 | 0.1 |
| | 50 | 0.0 | 0.05% | 0.000% | 49.7 | 0.0 | 0.0% | 0.2% | 0.0 | 0.2 |
| | 100 | 0.0 | 0.10% | 0.000% | 99.5 | 0.0 | 0.0% | 0.2% | 0.0 | 0.5 |
| | 250 | 0.0 | 0.25% | 0.000% | 248.7 | 0.0 | 0.0% | 0.2% | 0.0 | 1.2 |
| | 500 | 0.0 | 0.50% | 0.000% | 497.5 | 0.0 | 0.0% | 0.2% | 0.0 | 2.5 |
| | 1000 | 0.0 | 1.00% | 0.000% | 995.0 | 0.0 | 0.0% | 0.2% | 0.0 | 5.0 |
| 0.010% | 20 | 0.0 | 0.20% | 0.001% | 19.9 | 0.0 | 0.0% | 1.9% | 0.0 | 0.1 |
| | 50 | 0.0 | 0.50% | 0.001% | 49.7 | 0.0 | 0.0% | 1.9% | 0.0 | 0.2 |
| | 100 | 0.0 | 1.00% | 0.001% | 99.5 | 0.0 | 0.0% | 1.9% | 0.0 | 0.5 |
| | 250 | 0.0 | 2.47% | 0.001% | 248.7 | 0.0 | 0.1% | 1.9% | 0.0 | 1.2 |
| | 500 | 0.0 | 4.88% | 0.001% | 497.5 | 0.0 | 0.2% | 1.9% | 0.0 | 2.5 |
| | 1000 | 0.1 | 9.52% | 0.001% | 994.9 | 0.0 | 0.5% | 1.9% | 0.1 | 5.0 |
| 0.100% | 20 | 0.0 | 1.98% | 0.005% | 19.9 | 0.0 | 0.1% | 16.0% | 0.0 | 0.1 |
| | 50 | 0.0 | 4.88% | 0.005% | 49.7 | 0.0 | 0.2% | 16.0% | 0.0 | 0.2 |
| | 100 | 0.1 | 9.52% | 0.005% | 99.4 | 0.0 | 0.5% | 16.0% | 0.1 | 0.5 |
| | 250 | 0.2 | 22.13% | 0.005% | 248.5 | 0.0 | 1.2% | 16.0% | 0.2 | 1.2 |
| | 500 | 0.5 | 39.36% | 0.005% | 497.0 | 0.0 | 2.5% | 16.0% | 0.5 | 2.5 |
| | 1000 | 1.0 | 63.23% | 0.005% | 994.0 | 0.1 | 4.9% | 16.0% | 1.0 | 5.0 |

| | | No Screen | | Baseline Screening Test: 95% sensitivity, 99.5% specificity | | | | | | |
|---|---|---|---|---|---|---|---|---|---|---|
| | | | | Screen negatives out of NE "return to work" | | | | Screen positives out of NE "isolate/stay home" | | |
| **Pre-test Prevalence** | **Group Size NE** | **Number COVID19 Infected** | **Prob. at least 1 infected** | **Prevalence in screen negatives, 1 – NPV** | **Number TN** | **Number FN** | **Prob. at least 1 FN** | **Prevalence in screen positives, PPV** | **Number TP** | **Number FP** |
| 0.500% | 20 | 0.1 | 9.54% | 0.025% | 19.8 | 0.0 | 0.5% | 48.8% | 0.1 | 0.1 |
| | 50 | 0.2 | 22.17% | 0.025% | 49.5 | 0.0 | 1.2% | 48.8% | 0.2 | 0.2 |
| | 100 | 0.5 | 39.42% | 0.025% | 99.0 | 0.0 | 2.5% | 48.8% | 0.5 | 0.5 |
| | 250 | 1.2 | 71.44% | 0.025% | 247.5 | 0.1 | 6.1% | 48.8% | 1.2 | 1.2 |
| | 500 | 2.5 | 91.84% | 0.025% | 495.0 | 0.1 | 11.8% | 48.8% | 2.4 | 2.5 |
| | 1000 | 5.0 | 99.33% | 0.025% | 990.0 | 0.3 | 22.1% | 48.8% | 4.8 | 5.0 |
| 1.000% | 20 | 0.2 | 18.21% | 0.051% | 19.7 | 0.0 | 1.0% | 65.7% | 0.2 | 0.1 |
| | 50 | 0.5 | 39.50% | 0.051% | 49.3 | 0.0 | 2.5% | 65.7% | 0.5 | 0.2 |
| | 100 | 1.0 | 63.40% | 0.051% | 98.5 | 0.1 | 4.9% | 65.7% | 1.0 | 0.5 |
| | 250 | 2.5 | 91.89% | 0.051% | 246.3 | 0.1 | 11.8% | 65.7% | 2.4 | 1.2 |
| | 500 | 5.0 | 99.34% | 0.051% | 492.5 | 0.3 | 22.1% | 65.7% | 4.8 | 2.5 |
| | 1000 | 10.0 | 100.00% | 0.051% | 985.0 | 0.5 | 39.4% | 65.7% | 9.5 | 5.0 |
| 2.000% | 20 | 0.4 | 33.24% | 0.102% | 19.5 | 0.0 | 2.0% | 79.5% | 0.4 | 0.1 |
| | 50 | 1.0 | 63.58% | 0.102% | 48.8 | 0.1 | 4.9% | 79.5% | 1.0 | 0.2 |
| | 100 | 2.0 | 86.74% | 0.102% | 97.5 | 0.1 | 9.5% | 79.5% | 1.9 | 0.5 |
| | 250 | 5.0 | 99.36% | 0.102% | 243.8 | 0.3 | 22.1% | 79.5% | 4.8 | 1.2 |
| | 500 | 10.0 | 100.00% | 0.102% | 487.6 | 0.5 | 39.4% | 79.5% | 9.5 | 2.5 |
| | 1000 | 20.0 | 100.00% | 0.102% | 975.1 | 1.0 | 63.2% | 79.5% | 19.0 | 4.9 |

FN, false negative; FP, false positive; NPV, negative predictive value; PPV, positive predictive value; prob, probability; TN, true negative; TP, true positive.

Numbers in the table represent expected values, or what would be observed on average. Actual numbers will vary.

**Supplementary Table 8. Impact of the 1-stage sample pooling approach for pool sizes 5 or 10 versus an individual testing approach (not pooled) using a test with individual sensitivity of 80% and specificity of 99.5%.**

| | | Pool Parameters | | | Tests | | Declared individual results negative out of NE "return to work" | | | | Declared individual results positive out of NE "isolate/stay home" | | | |
|---|---|---|---|---|---|---|---|---|---|---|---|---|---|---|
| **Pre-test prev** | **NE** | **k** | **Pool prev** | **Pool sens** | **#** | **%** | **Prev. in screen neg** | **TN** | **FN** | **Prob at least 1 FN** | **Prev in screen pos** | **TP** | **FP** | **% no pos*** |
| **0.01%** | 1000 | Not pooled | | | 1000 | 100% | 0.00% | 994.9 | 0.0 | 1.9% | 1.6% | 0.1 | 5.0 | 1% |
| | 1000 | 5 | 0.05% | 78.5% | 200 | 20% | 0.00% | 994.7 | 0.0 | 2.1% | 1.5% | 0.0 | 5.3 | 34% |
| | 1000 | 10 | 0.10% | 77.5% | 100 | 10% | 0.00% | 994.3 | 0.0 | 2.0% | 1.3% | 0.1 | 5.6 | 34% |
| **0.1%** | 1000 | Not pooled | | | 1000 | 100% | 0.02% | 994.0 | 0.2 | 17.7% | 13.7% | 0.8 | 5.0 | 0% |
| | 1000 | 5 | 0.50% | 78.5% | 200 | 20% | 0.02% | 990.8 | 0.2 | 18.0% | 8.9% | 0.8 | 8.2 | 17% |
| | 1000 | 10 | 1.00% | 77.5% | 100 | 10% | 0.02% | 987.0 | 0.2 | 19.4% | 6.2% | 0.8 | 12.0 | 17% |
| **1.0%** | 1000 | Not pooled | | | 1000 | 100% | 0.20% | 985.0 | 2.0 | 86.2% | 61.7% | 8.0 | 5.0 | 0% |
| | 1000 | 5 | 4.90% | 78.5% | 200 | 20% | 0.21% | 954.1 | 2.0 | 86.1% | 18.2% | 8.0 | 35.9 | 0% |
| | 1000 | 10 | 9.56% | 77.5% | 100 | 10% | 0.23% | 917.8 | 2.2 | 87.8% | 9.9% | 7.9 | 72.1 | 0% |

FN, NE with false negative results; FP, NE with false positive results; k, pool size; Neg, negative; pos, positive; prev, prevalence; prob, probability; sens, sensitivity; TN, NE with true negative results; TP, NE with true positive results; #, number; %, percent.

*Percentage of simulations with no (zero) positive test results.

The grey shaded rows represent individual sample testing (no pooling). Numbers in the table represent average values across 10,000 sets of simulated values, or what would be observed on average.  Actual numbers will vary.

**Supplementary Table 9. Impact of the Dorfman 2-stage sample pooling approach for pool sizes 5 or 10 versus an individual testing approach (not pooled/retested) using a test with individual sensitivity of 80% and specificity of 99.5%.**

| | | Pool Parameters | | | Tests | | Declared individual results negative out of NE "return to work" | | | | Declared individual results positive out of NE "isolate/stay home" | | | |
|---|---|---|---|---|---|---|---|---|---|---|---|---|---|---|
| **Pre-test prev** | **NE** | **k** | **Pool prev** | **Pool sens** | **#** | **%** | **Prev in screen neg** | **TN** | **FN** | **Prob at least 1 FN** | **Prev in screen pos** | **TP** | **FP** | **% no pos*** |
| **0.01%** | 1000 | Not pooled/retested | | | 1000 | 100% | 0.00% | 994.9 | 0.0 | 1.9% | 1.6% | 0.1 | 5.0 | 1% |
| | 1000 | 5 | 0.05% | 78.5% | 205 | 21% | 0.00% | 999.9 | 0.0 | 3.5% | 70.6% | 0.1 | 0.0 | 91% |
| | 1000 | 10 | 0.10% | 77.5% | 106 | 11% | 0.00% | 999.9 | 0.0 | 3.5% | 69.6% | 0.1 | 0.0 | 91% |
| **0.1%** | 1000 | Not pooled/retested | | | 1000 | 100% | 0.02% | 994.0 | 0.2 | 17.7% | 13.7% | 0.8 | 5.0 | 0% |
| | 1000 | 5 | 0.50% | 78.5% | 209 | 21% | 0.04% | 998.9 | 0.4 | 29.8% | 95.1% | 0.7 | 0.0 | 51% |
| | 1000 | 10 | 1.00% | 77.5% | 113 | 11% | 0.04% | 998.9 | 0.4 | 30.6% | 93.2% | 0.6 | 0.1 | 51% |
| **1.0%** | 1000 | Not pooled/retested | | | 1000 | 100% | 0.20% | 985.0 | 2.0 | 86.2% | 61.7% | 8.0 | 5.0 | 0% |
| | 1000 | 5 | 4.90% | 78.5% | 244 | 24% | 0.36% | 989.8 | 3.6 | 97.4% | 97.6% | 6.4 | 0.2 | 0% |
| | 1000 | 10 | 9.56% | 77.5% | 180 | 18% | 0.38% | 989.6 | 3.7 | 97.5% | 95.1% | 6.3 | 0.4 | 0% |

FN, NE with false negative results; FP, NE with false positive results; k, pool size; Neg, negative; pos, positive; prev, prevalence; prob, probability; sens, sensitivity; TN, NE with true negative results; TP, NE with true positive results; #, number; %, percent.

*Percentage of simulations with no (zero) positive test results.

The grey shaded rows represent individual sample testing (no pooling and no retesting). Numbers in the table represent average values across 10,000 sets of simulated values, or what would be observed on average. Actual numbers will vary.

**Supplementary Table 10. Expected number of days and percent of 14-day contagious period that an infected contagious employee remains on site until detected as test positive with a 1- or 2-day turnaround time**

| | 1-day turnaround time | | | | | 2-day turnaround time | | | | |
|---|---|---|---|---|---|---|---|---|---|---|
| Testing Interval (days) | No. tests per TINF | 60% Sens | 70% Sens | 80% Sens | 90% Sens | No. tests per TINF | 60% Sens | 70% Sens | 80% Sens | 90% Sens |
| 1 | 14 | 1.7 (12%) | 1.4 (10%) | 1.2 (9%) | 1.1 (8%) | | | | | |
| 2 | 7 | 2.3 (17%) | 1.9 (13%) | 1.5 (11%) | 1.2 (9%) | 7 | 3.3 (24%) | 2.9 (20%) | 2.5 (18%) | 2.2 (16%) |
| 4 | 4 | 3.5 (25%) | 2.7 (19%) | 2.0 (14%) | 1.4 (10%) | 4 | 4.5 (32%) | 3.7 (26%) | 3.0 (21%) | 2.4 (17%) |
| 7 | 2 | 4.8 (34%) | 3.6 (26%) | 2.6 (19%) | 1.8 (13%) | 2 | 5.6 (40%) | 4.6 (33%) | 3.6 (26%) | 2.8 (20%) |
| 14 | 1 | 6.2 (44%) | 4.9 (35%) | 3.6 (26%) | 2.3 (16%) | 1 | 6.8 (49%) | 5.6 (40%) | 4.4 (31%) | 3.2 (23%) |

TINF, time infected, asymptomatic but contagious; sens, sensitivity.

Numbers in the table represent expected values, or what would be observed on average. Actual numbers will vary.

**Supplementary Table 11. Expected number of days and percent of 21-day contagious period that an infected contagious employee remains on site until detected as test positive with a 1- or 2-day turnaround time**

| | 1-day turnaround time | | | | | 2-day turnaround time | | | | |
|---|---|---|---|---|---|---|---|---|---|---|
| Testing Interval (days) | No. tests per TINF | 60% Sens | 70% Sens | 80% Sens | 90% Sens | No. tests per TINF | 60% Sens | 70% Sens | 80% Sens | 90% Sens |
| 1 | 21 | 1.7 (8%) | 1.4 (7%) | 1.2 (6%) | 1.1 (5%) | | | | | |
| 2 | 11 | 2.3 (11%) | 1.9 (9%) | 1.5 (7%) | 1.2 (6%) | 11 | 3.3 (16%) | 2.9 (14%) | 2.5 (12%) | 2.2 (11%) |
| 4 | 6 | 3.6 (17%) | 2.7 (13%) | 2.0 (10%) | 1.4 (7%) | 6 | 4.6 (22%) | 3.7 (18%) | 3.0 (14%) | 2.4 (12%) |
| 7 | 3 | 5.3 (25%) | 3.9 (19%) | 2.7 (13%) | 1.8 (8%) | 3 | 6.2 (30%) | 4.9 (23%) | 3.7 (18%) | 2.8 (13%) |
| 14 | 2 | 7.6 (36%) | 5.7 (27%) | 4.0 (19%) | 2.5 (12%) | 2 | 8.4 (40%) | 6.7 (32%) | 5.0 (24%) | 3.4 (16%) |

TINF, time infected, asymptomatic but contagious; sens, sensitivity

Numbers in the table represent expected values, or what would be observed on average. Actual numbers will vary.

$$TC = (TA)\,se + (TA+TI)(1-se)se + (TA+2TI)(1-se)^2 se + \cdots + (TA+(m-1)TI)(1-se)^{m-1} se + TINF(1-se)^m$$

Probability of testing positive × time to test result

Probability of testing negative **once** then testing positive × time to test result at next test interval

Probability of testing negative **twice** then testing positive × time to test result at next test interval

Probability of testing negative **all m times** during contagious time (resolved & never detected by testing)

**Supplementary Figure 1. Formula for calculating the expected number of days an infected employee will be on site before being detected**

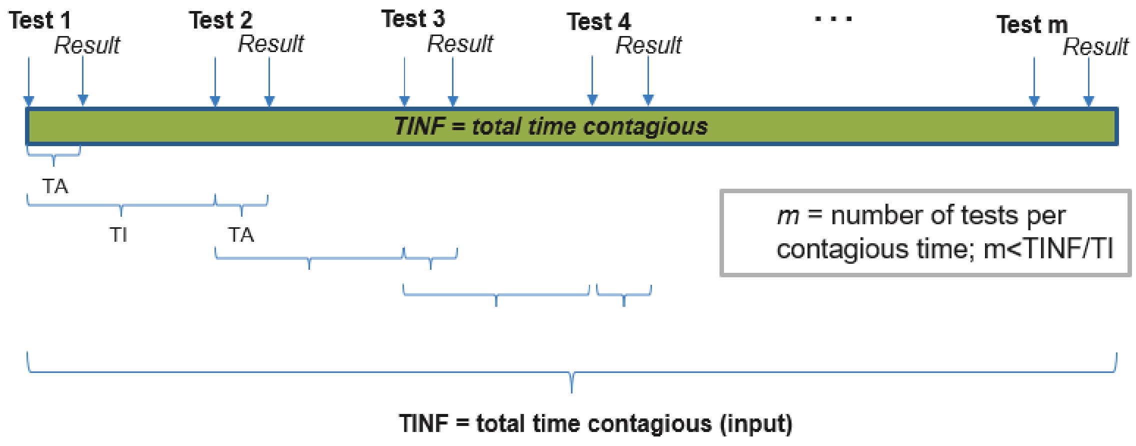


**Supplementary Figure 2. Description of testing and timing of results in relation to TA, TI and TINF to calculate the expected number of days an infected employee will be on site before being detected**
